# Speech disturbances in schizophrenia: assessing cross-linguistic generalizability of NLP automated measures of coherence

**DOI:** 10.1101/2022.03.28.22272995

**Authors:** Alberto Parola, Jessica Mary Lin, Arndis Simonsen, Vibeke Bliksted, Yuan Zhou, Huiling Wang, Lana Inoue, Katja Koelkebeck, Riccardo Fusaroli

## Abstract

**Introduction:** Language disorders – disorganized and incoherent speech in particular - are distinctive features of schizophrenia. Natural language processing (NLP) offers automated measures of incoherent speech as promising markers for schizophrenia. However, the scientific and clinical impact of NLP markers depends on their generalizability across contexts, samples, and languages, which we systematically assessed in the present study relying on a large, novel, cross-linguistic corpus.

**Methods:** We collected a Danish (DK), German (GE), and Chinese (CH) cross-linguistic dataset involving transcripts from 187 participants with schizophrenia (111DK, 25GE, 51CH) and 200 matched controls (129DK, 29GE, 42CH) performing the Animated Triangle task. Fourteen previously published NLP coherence measures were calculated, and between-groups differences and association with symptoms were tested for cross-linguistic generalizability.

**Results:** One coherence measure robustly generalized across samples and languages. We found several language-specific effects, some of which partially replicated previous findings (lower coherence in German and Chinese patients), while others did not (higher coherence in Danish patients). We found several associations between symptoms and measures of coherence, but the effects were generally inconsistent across languages and rating scales.

**Conclusions:** Using a cumulative approach, we have shown that NLP findings of reduced semantic coherence in schizophrenia have limited generalizability across different languages, samples, and measures. We argue that several factors such as sociodemographic and clinical heterogeneity, cross-linguistic variation, and the different NLP measures reflecting different clinical aspects may be responsible for this variability. Future studies should take this variability into account in order to develop effective clinical applications targeting different patient populations.

## Introduction

Language disturbances have been a hallmark of schizophrenia since the first definitions of the disorder (Bleuler, 1911; Kraepelin, 1919). They are particularly evident at the discourse level - ranging from reduced syntactic complexity to loss of semantic coherence and cohesion -, and are often associated with specific symptoms (e.g., formal thought disorders). Language disorders can seriously impair the patients’ social functioning and communicative ability (e.g., Bliksted et al., 2014; Green et al., 2015; Gallagher & Varga; 2015; Parola et al., 2018; 2021) and are pervasive: they are an early distinctive feature of schizophrenia, preceding the onset of initial psychosis, and occur in individuals at high clinical risk as well as in patients’ relatives (Bedi et al., 2015; Corcoran et al., 2018; Rezaii et al., 2019). Therefore, language disorders - and disorganized and incoherent speech in particular - could play a critical role for developing digital phenotyping of schizophrenia (Corcoran et al., 2020; de Boer et al., 2018; De Boer et al., 2020; Hitczenko et al., 2021).

Recent advances in natural language processing (NLP) techniques - e.g., topic modeling (Rezaii et al., 2019), word embeddings (Bedi et al., 2015; Corcoran et al., 2018; Elvevåg et al., 2007; Holshausen et al., 2014; Just et al., 2020; Tang et al., 2021; Voppel et al., 2021), speech graph analysis (Mota et al., 2014, 2017), and semantic density quantification (Rezaii et al., 2019) - could provide quantitative, cost-effective, and automated measures of incoherent speech. Indeed, automated analyses of linguistic content and coherence have variously found lower coherence and semantic density in schizophrenia, and individuals at high clinical risk.

These findings suggest that NLP techniques may complement clinical observations and constitute a window into the social and emotional features of the disorder (Cohen et al., 2021; Corcoran & Cecchi, 2020). However, a critical obstacle to any concrete use of these findings is that it is not clear whether the findings would replicate and generalize to new samples and populations, an overarching problem for clinical and social sciences (Hitczenko et al., 2021; Parola et al., 2020; Rocca & Yarkoni, 2021; Rybner et al., 2021). Indeed, a closer look reveals clearly contradictory results: linguistic measures are inconsistently associated with symptoms, and findings vary across different rating scales and samples (Bedi et al., 2015; Corcoran et al., 2018; Haas et al., 2020; Morgan et al., 2021; Pauselli et al., 2018; Sarzynska-Wawer et al., 2021; Tang et al., 2021).

Such inconsistencies may have several causes. Sample sizes are usually small (median size for participants with schizophrenia = 34.5), and given the heterogeneity of schizophrenia, differences in demographic and clinical characteristics between studies can be quite large, and findings be overfit to the specific sample. Moreover, linguistic and/or cultural specificity may seriously affect coherence patterns, thus leading to differences in language impairments between samples collected in different countries, as well as differences in their specific association with symptomatology (Palaniyappan, 2021; Sumiyoshi et al., 2004, 2014; Wydell & Butterworth, 1999). For example, native speakers of different languages display differences in word retrieval and word processing strategies and in the use of pauses, hesitations and false starts (Sumiyoshi et al., 2004; Ishkhanyan et al., 2020; Palaniyappan, 2020). Thus it is not clear how well the findings of previous studies can generalize to different linguistic and/or cultural groups. Finally, automated measures of linguistic content have been operationalized in very different ways and differ substantially across studies-potentially reflecting different psychopathological dimensions - and these differences may account for the differences in findings.

To move beyond this situation, this study showcases a cumulative scientific approach capable of systematically assessing the impact of previous findings on current data and integrating the new findings into a global framework. Such a framework promotes the systematic assessment of previous findings and different automated measures of coherence across contexts and samples with different clinical, demographic, cultural and linguistic profiles (Corcoran et al., 2018; Fusaroli et al., 2021; Rybner et al., 2021). This will provide a more robust predictive performance assessment, but it may also provide more reliable foundations for theory development (e.g. generative modeling of incoherence) and accordingly improved understanding of language disturbances (Press et al., 2022; Rocca & Yarkoni, 2021).

First, we systematically reviewed the literature to identify replicable automated NLP coherence measures that characterize the language of patients with schizophrenia, and effect sizes of previous findings for critical comparison. Second, we assembled a large cross-linguistic (Danish, German, Chinese) corpus consisting of multiple speech transcriptions from patients with schizophrenia and matched controls. Third, we critically and systematically assessed how well previous findings generalized to the new corpus and were robust to language and sample variations. By comparing literature-informed (Brand et al., 2019; Fusaroli et al., 2021) to more traditional analyses (relying on regularizing priors), we can more directly assess how the data supports or strays from previously found patterns. Fourth, we provided a more systematic evaluation of the heterogeneities involved in the study. We explicitly model heterogeneity by assessing the associations between measures of coherence and clinical ratings of psychopathology. Crucially, we were also able to estimate the robustness of coherence measures, within and between subjects, samples and languages, as well as their variability. Finally, we adopt an approach where we rely on open source software, extract features in a reproducible manner using openly available scripts, carefully describe the methodology used, and test the robustness of the results to variations in methodology.

The aim of the study is to directly address the problem of generalizability of previous NLP results and to develop a critical and systematic approach that can further the understanding of language disorders and their relationship with symptoms in schizophrenia.

## Methods

### Participants

We collected a Danish (DK), German (GE), and Chinese (CH) cross-linguistic dataset involving 187 participants with schizophrenia(111 DK, 25 GE, 51 CH) and 200 matched controls (HC) (129 DK, 29 GE, 42 CH). The samples for the present study were collected in separate studies assessing mentalizing ability in patients with schizophrenia and healthy controls. Information on demographics, IQ, psychopathology, and social functioning is summarized in **Table 1**. Detailed information on each study is reported in the **Supplementary Material 1 (SM1)**.

**Table 1.** Demographic and clinical characteristics of patients with schizophrenia (SCZ) and healthy controls (HC).

| Corpus | Danish |  | German |  | Chinese |  |
| --- | --- | --- | --- | --- | --- | --- |
| **Diagnosis** | SCZ<br>N = 111 | HC<br>N = 129 | SCZ<br>N = 25 | HC<br>N = 29 | SCZ<br>N = 51 | HC<br>N = 42 |
| **N. of transcripts** | N = 944 | N = 1102 | N = 298 | N = 346 | N = 406 | N = 321 |
| **Age** | 26.9<br>(9.57) | 26.4<br>(8.78) | 29.2<br>(8.48) | 30.7<br>(7.56) | 27.2 (7.22) | 28.5<br>(7.72) |
| **Education** | 12.8<br>(2.76) | 14.8<br>(2.54) | 12.1<br>(1.48) | 12.3<br>(1.11) | 12.7 (2.69) | 14.4<br>(2.12) |
| **Gender (n. of females and %)** | 46<br>(41%) | 59<br>(46%) | 11<br>(44%) | 12<br>(41%) | 23<br>(45%) | 18<br>(43%) |
| **Verbal IQ** | 89.12 (18.67) | 102.02 (15.94) | NA | NA | 96.03 (16.53) | 101.72 (14.17) |
| **SANS total** | 9.70<br>(4.39) | NA | NA | NA | 7.61 (3.01) | NA |
| **SAPS total** | 10.38 (4.90) | NA | NA | NA | 7.16 (4.79) | NA |
| **PANSS total** | NA | NA | 52.87 | 10.09 | 75.72 | 10.46 |
| **PANSS negative** | NA | NA | 14.12 | 3.90 | 20.01 | 5.25 |
| **PANSS positive** | NA | NA | 10.64 | 2.72 | 18.54 | 4.36 |
| **Illness duration (months)** | 8.89<br>(6.70) | NA | 28.96 (47.42) | NA | 62.97 (68.20) | NA |

### Speech samples

Speech samples were collected using the Animated Triangles task(Abell et al., 2000; Castelli et al., 2000). The task is used to measure theory of mind (ToM), and it consists of twelve video clips representing an interaction between animated triangles. In the four random clips the two triangles are moving randomly and unintentionally (e.g., bouncing about), in the four ToM clips the triangles are interacting intentionally (e.g., the large triangle trying to convince the small triangle to come outside), and in the other four clips they are merely performing an activity alone or together. The duration of each animation is approximately 40 seconds. The participants were asked to provide an interpretation of what was going on in each animation and their answers were audio-recorded and then transcribed by research assistants (see **SM2** for more details on the task). After the transcription, fillers such as ‘uhm’ and “ehm” (see **SM3** for more details) were removed. No other preprocessing was performed; specifically, interjections were not removed.

### Speech pre-processing

We prepared the data for NLP-based analysis by using the UDPipe Natural Language Processing - Text Annotation in R (Straka et al., 2016). First, words were tokenized (identified as parts of speech), and then each transcript was parsed into phrases, using rules of grammar for each specific language. Words were then converted to the roots from which they are inflected, or lemmatized. The resulting pre-processed speech data yielded for each transcript a series of lemmatized words, maintaining the original order in which they were spoken. We then used fastText pre-trained models for the different languages (Bojanowski et al., 2017) to vectorize the tokenized speech samples, yielding a 300-dimensional vector for each word. After that, we computed semantic coherence between words, i.e., word-to-word similarity, by calculating the cosine similarity between the corresponding vectors associated with each word. The cosine similarity values range between − 1 and 1, with − 1 representing the lowest similarity and 1 the highest similarity between two words (see **SM3** for more details).

### Literature search and NLP measures of coherence

We systematically screened the current literature - following the indications of the Preferred Reporting Items for Systematic Reviews and Meta-Analyses Guidelines (PRISMA) statement (Rethlefsen et al., 2021) - to identify previous studies in the literature which quantified semantic coherence in schizophrenia using NLP automated methods (full details on the search on **SM4**). Among the final set of selected studies, we selected scalable measures of coherence, i.e. easier to apply to a larger set of languages (without ad hoc wordlists, etc), and to corpora with limited size (e.g. excluding training deep learning methods on the corpus). We found 14 studies using 14 different NLP measures to quantify semantic coherence in schizophrenia: we thus derived those coherence measures and tried to replicate their results on our corpus (**Table 2** and **SM4**). Median and interquartile range (IQR) were calculated for each of these measures (see **SM4**). We report in the main manuscript only results for median, while IQR results are reported in **SM5**. We opted to use median and IQR of coherence measures, even if previous studies used a wide variety of descriptors (e.g. standard deviation, maximum, minimum, etc..) because they are more robust to measurement errors. Note that any difference (e.g. in data preprocessing, or word embeddings employed) between the original studies and the current one is motivated by the goal of building a data analysis framework able to extract different measures of coherence and compare them in a scalable way across different samples and languages, and fully detailed in **SM4**.

**Table 2.**
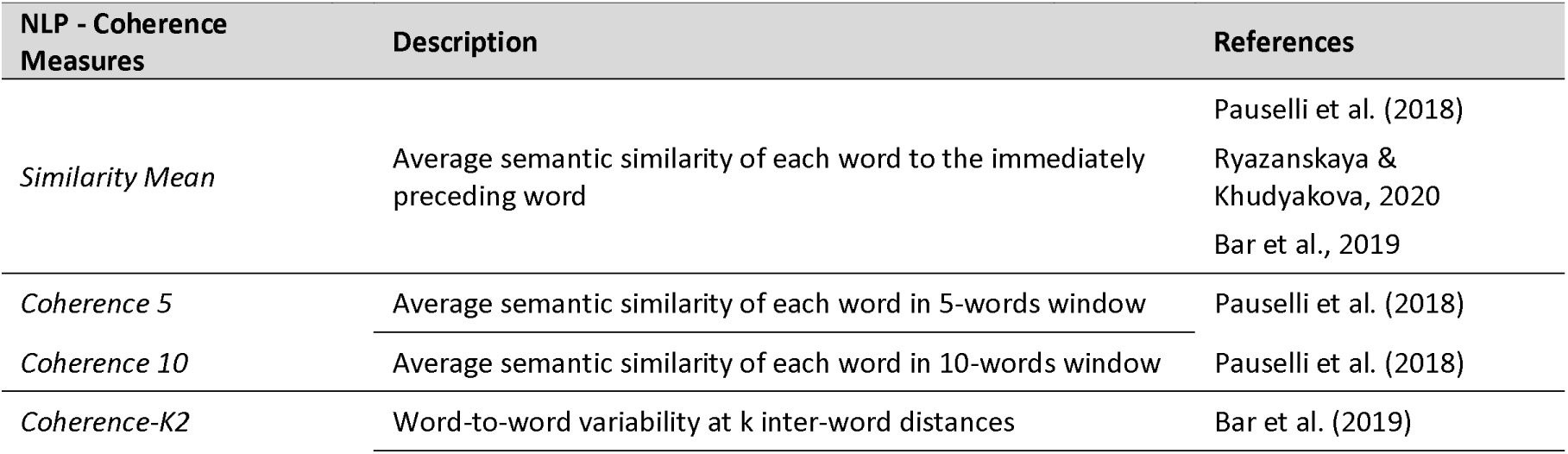
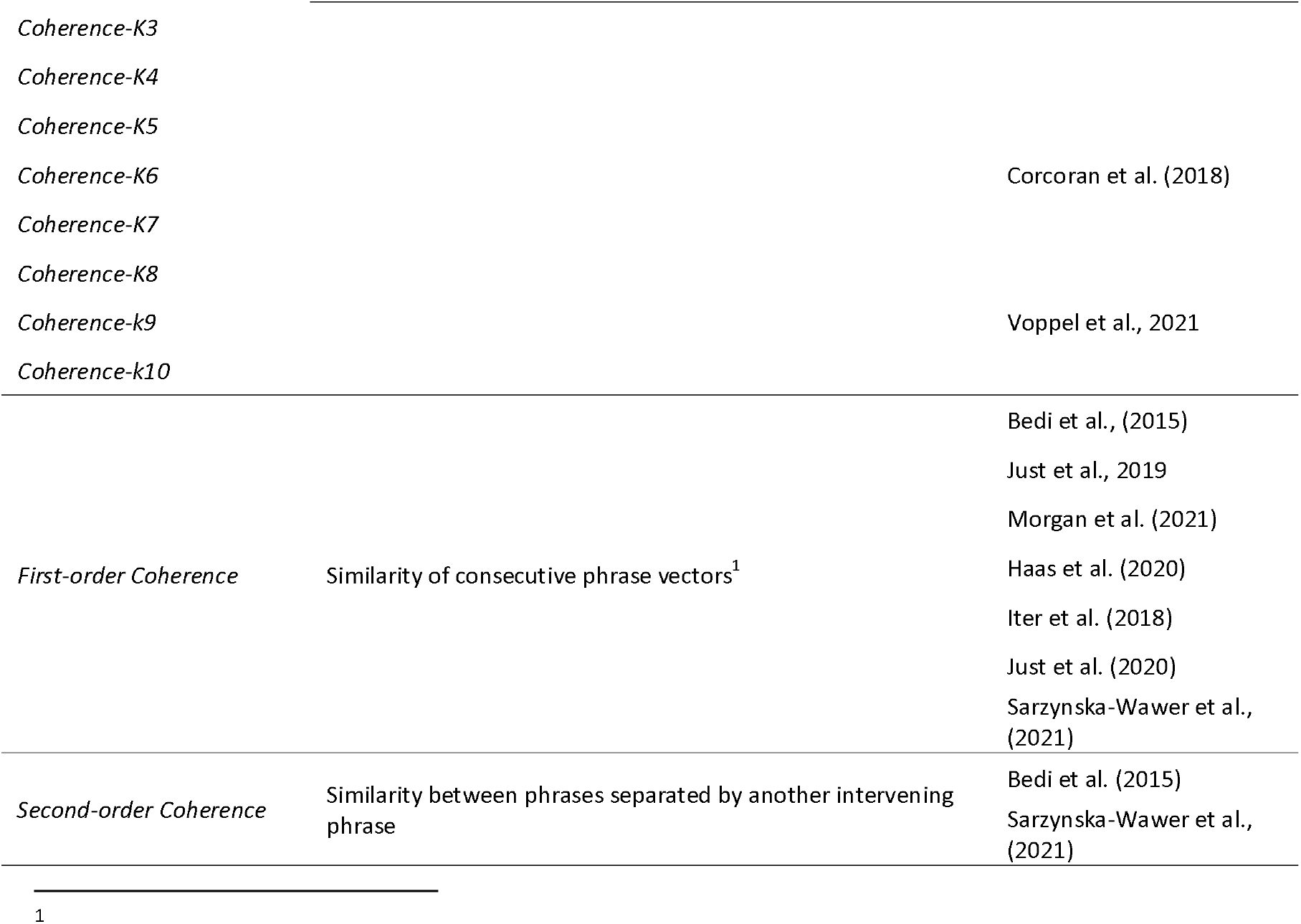
NLP coherence measures identified in the previous studies and derived in the present research.

### Analysis of effect of diagnosis on (differences in) coherence measures

To estimate the differences between individuals with schizophrenia and HC in the different coherence measures, we used Bayesian multilevel regression models on the current data with each coherence measure as outcome, and diagnosis (schizophrenia vs. HC) and language (DK, GE, CH) as predictors. Within the same model, we separately assessed the effect of diagnosis for each language, and modeled varying effects of participants, i.e., intercepts and slopes, separately for each group and language. For each coherence measure, we built a model with weakly informative priors, i.e., expectations of no effects of diagnosis, thus conservatively regularizing the model parameters, reducing overfitting and leading to improved predictions (Gelman et al., 2020). We then built a second model with informed priors (when available), that is summary effect sizes (ES, see **SM5**), and compared results across the two models. We aimed to assess whether the effects of diagnosis are robust across changes of priors, and whether the skeptical or informed priors led to more robust inference, that is, in lower estimated out-of-sample error - measured in terms of Leave-One-Out based stacking weights. To evaluate the potential role of gender (male vs. female), age and level of intelligence we built additional models, one per each moderator interacting with group separately in the three languages. We then reported the model estimates for the interaction, including credible (i.e., Bayesian confidence) intervals (CIs) and evidence ratios (ERs), i.e. evidence in favor of the effect observed against alternative hypotheses. When ER was weak (below 10, that is, less than ten times as much evidence for the effect as for alternative hypotheses), we also calculated the Evidence Ratio in favor of the null hypothesis. Further details are presented in the **SM5**. Note also that we report additional analyses in the **SM6** to assess the robustness of the findings: we repeated all analyses by: 1) including fillers and 2) including fillers and punctuation 3) explicitly assessing the association between transcript length (total number of words) and the different coherence measures. The results support our main findings and we report in the manuscript only qualitative divergences.

### Analysis of the relationship between coherence measures and clinical ratings

To assess the relationship between the coherence measures and clinical ratings, we built Bayesian multilevel regression models with each coherence measure as outcome, and clinical features (one at a time) as ordinal predictors. We separately assessed the relationship between the different coherence measures and clinical ratings for each language, and modeled varying effects of participants, i.e. intercepts and slopes, separately for each language. This analysis was performed on the schizophrenia group only (see **SM5** for more details). All the code used for the analysis and the extracted features are openly available (see **SM7**).

## Results

### Effect of Diagnosis

The detailed results are reported in **Table 2** and **Figure 1**. We only partially replicated previous findings: reduced Similarity mean, Coherence 10, and Coherence-K in schizophrenia were found only in the Chinese and German corpus, while increased Coherence-K and increased Coherence 10 measures were found in the Danish corpus. Reduced first order coherence in schizophrenia was found in the Danish and Chinese corpus, while reduced second-order coherence was found in all the corpus. We found a lower total number of words in schizophrenia in the Danish and Chinese corpus. Globally, we found important differences within (i.e. between the diverse coherence measures) and between languages. In agreement with the inconsistent replications, the informed models were more robust and generalizable to new data (LOO weights for informed models above .75) in less than half of the models (3 on 8, in 1 model there was no difference), indicating that prior findings were not fully representative of the current samples. Gender, age, education and level of intelligence of the participants also affected the group differences, although inconsistently across languages. Our results are robust to the inclusion of fillers and punctuations in the analysis, even if some differences are present and discussed in **SM6**. We also found a positive association between transcript length and the various coherence measures, that is longer transcripts tend to have higher coherence values.

**Table 2.**
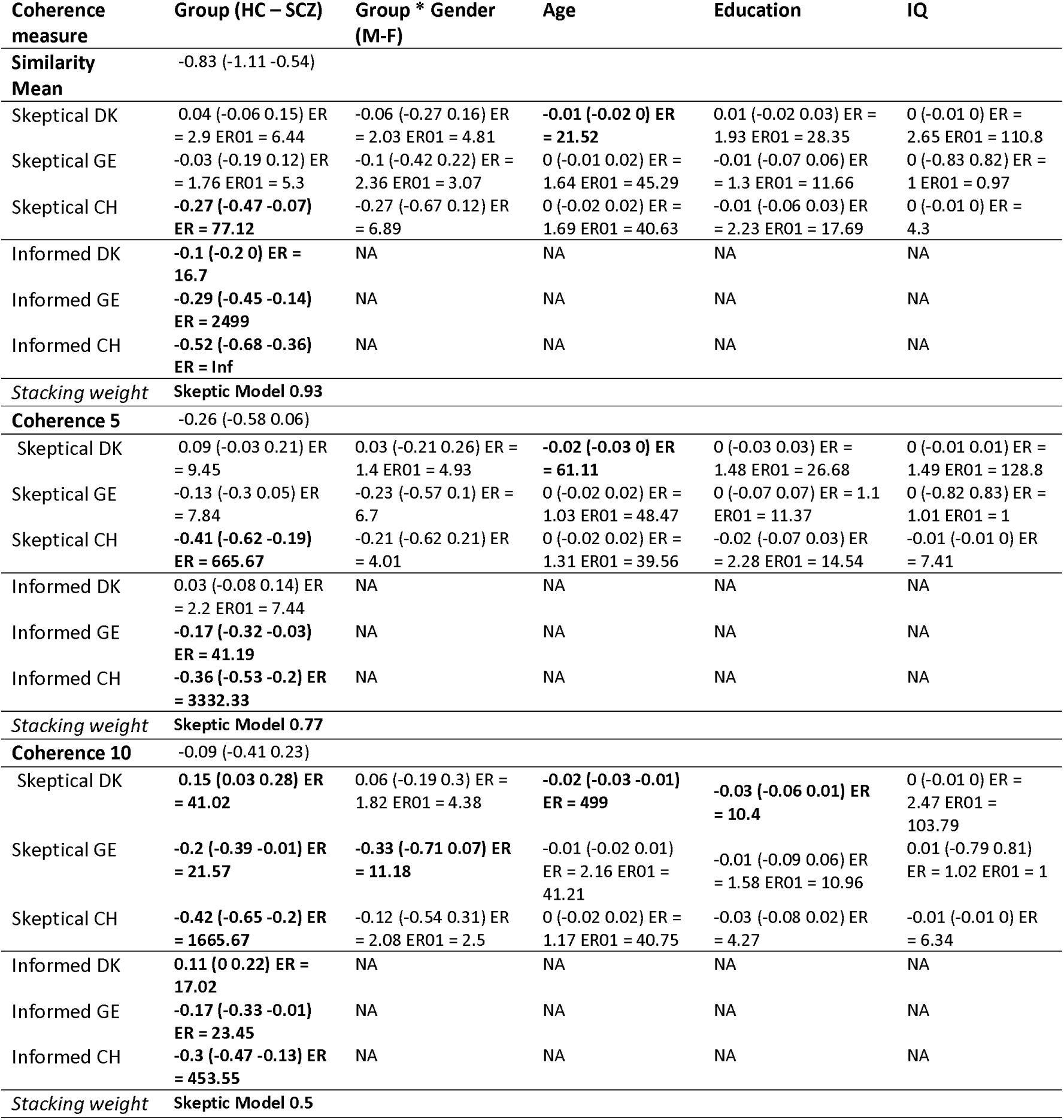
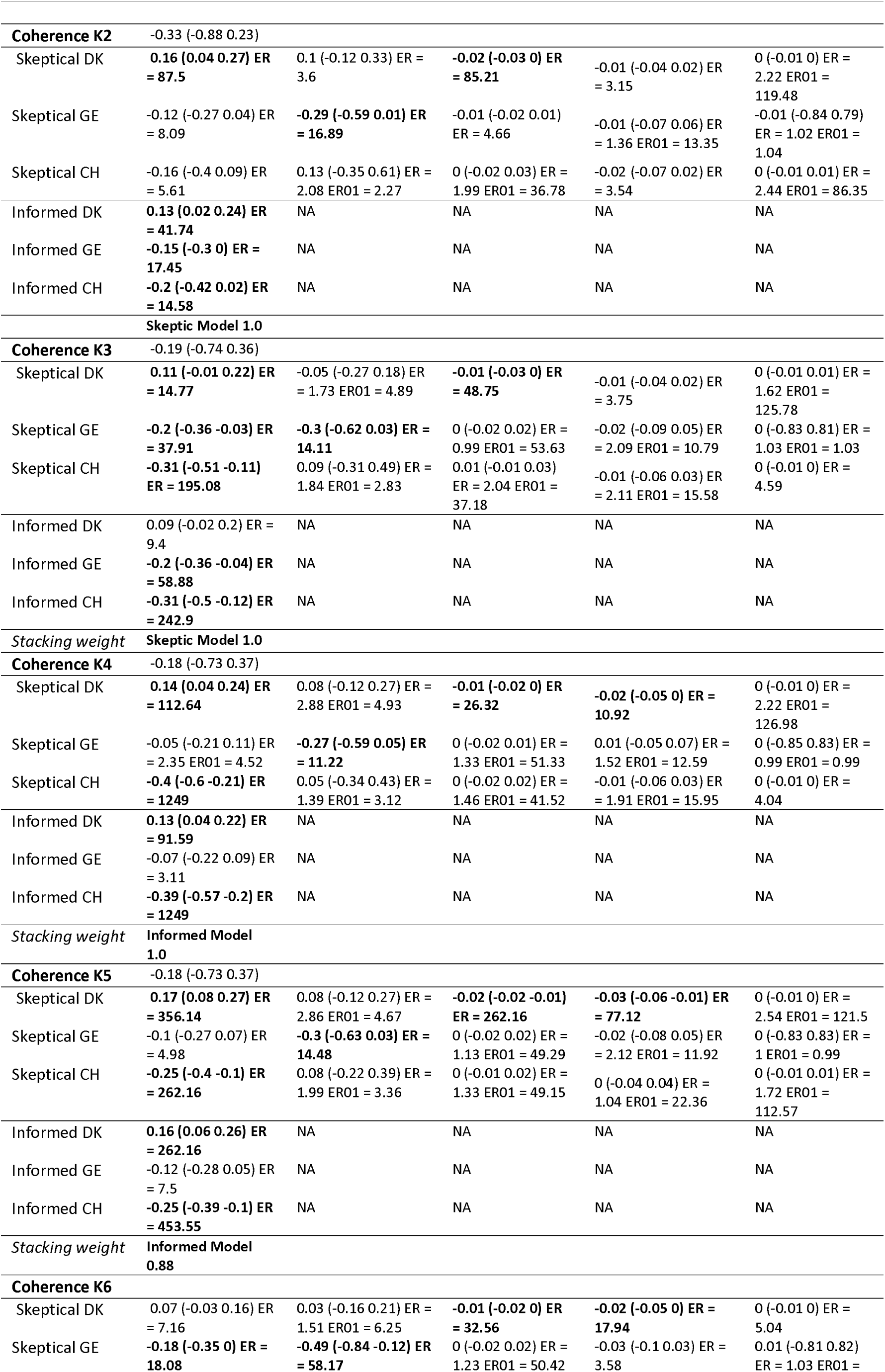
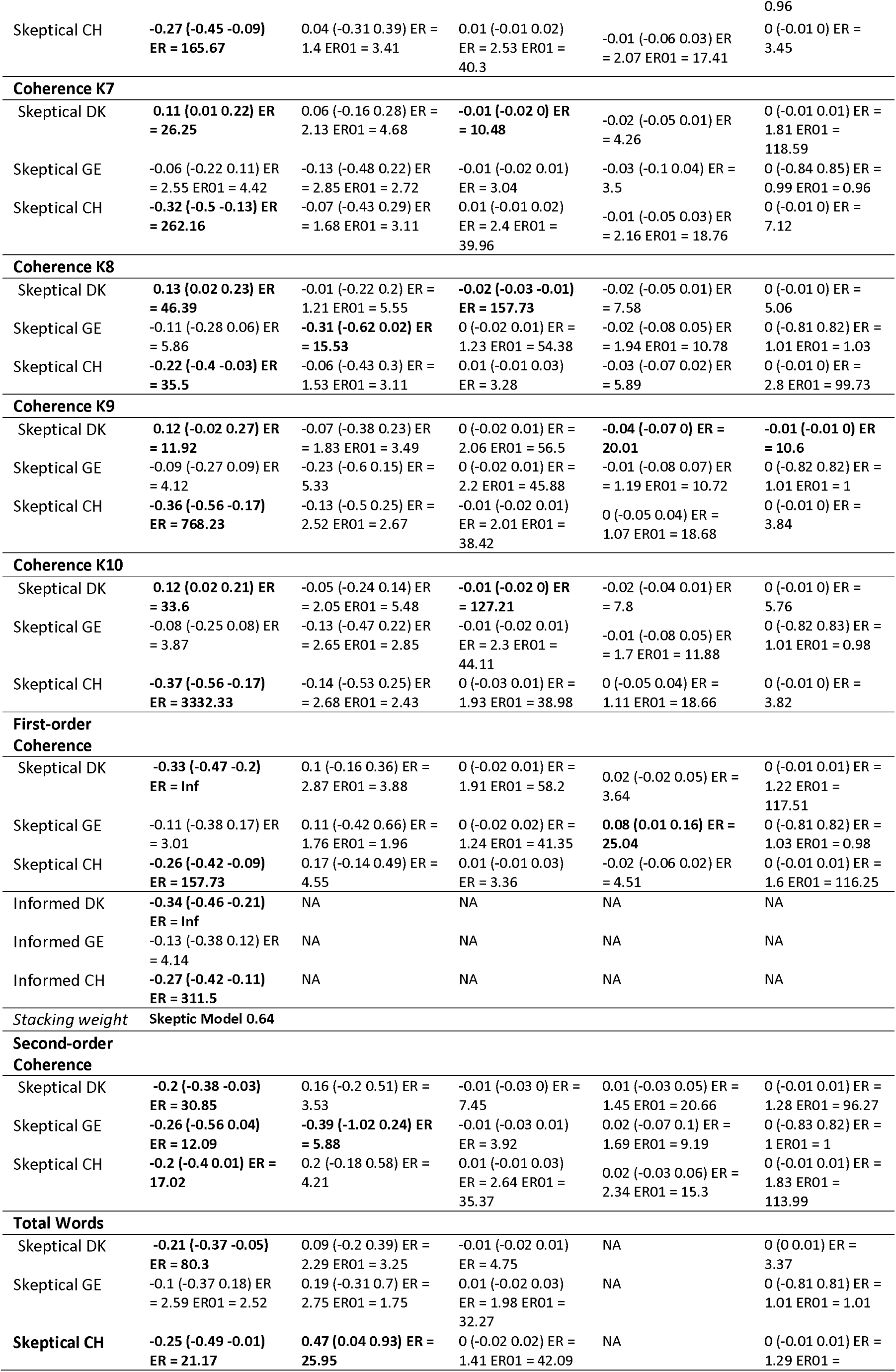
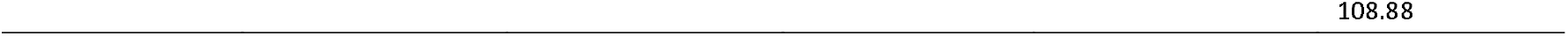
Estimated standardized mean difference (controls – patients with schizophrenia) for the fourteen coherence measures, as estimated separately by the informed and the skeptical models.

**Figure 1.**
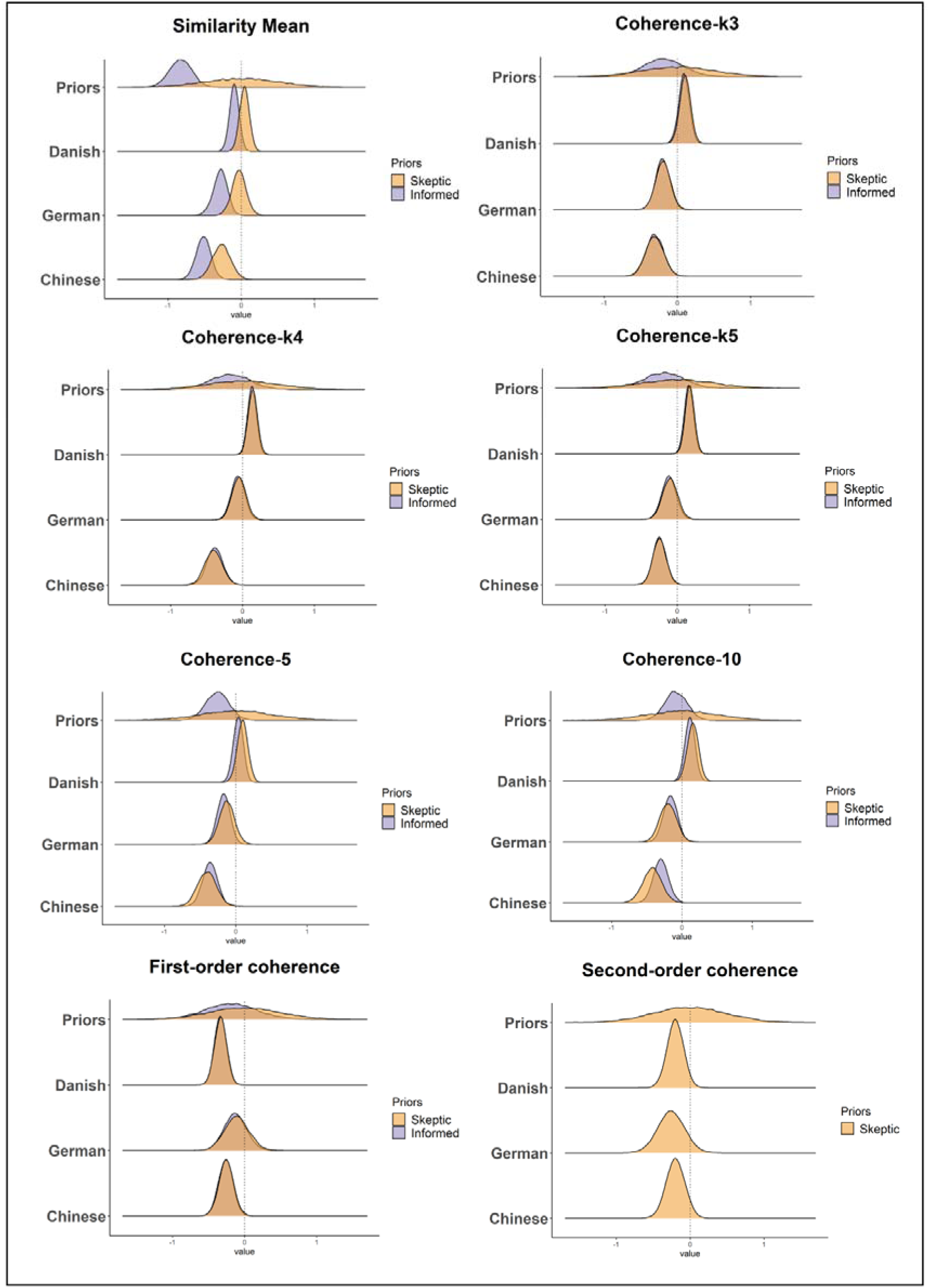
Comparing informed and skeptical expectations and results. Each panel presents a separate coherence measure, with the x-axis corresponding to standardized mean differences (schizophrenia - HC) equivalent to Hedges’ g, with estimates above 0 indicating higher scores for patients with schizophrenia.

### Effect of Symptoms

Detailed results are reported in **Table 4, Table 5 and Table SM5_B**. Globally, clinical ratings of symptoms correlate with NLP measures of semantic coherence. We found several associations between Coherence-k, Coherence 5-10 and First-order coherence measures, and SAPS ratings (Global SAPS, SAPS FTD, Derailment, Tangentiality, Incoherence). However, these associations were not robust across languages, and the direction of the effects were often inconsistent. We found more consistent associations SANS ratings and several coherence measures, i.e. higher SANS ratings (global SANS, global alogia, poverty of content and poverty of speech) were generally associated with reduced coherence. We found a very weak association between PANSS symptoms and coherence measures (see **Table SM5**). Most of the correlations were between small and moderate (5-20% of explained variance), and varied across languages and rating scales.

**Table 4.**
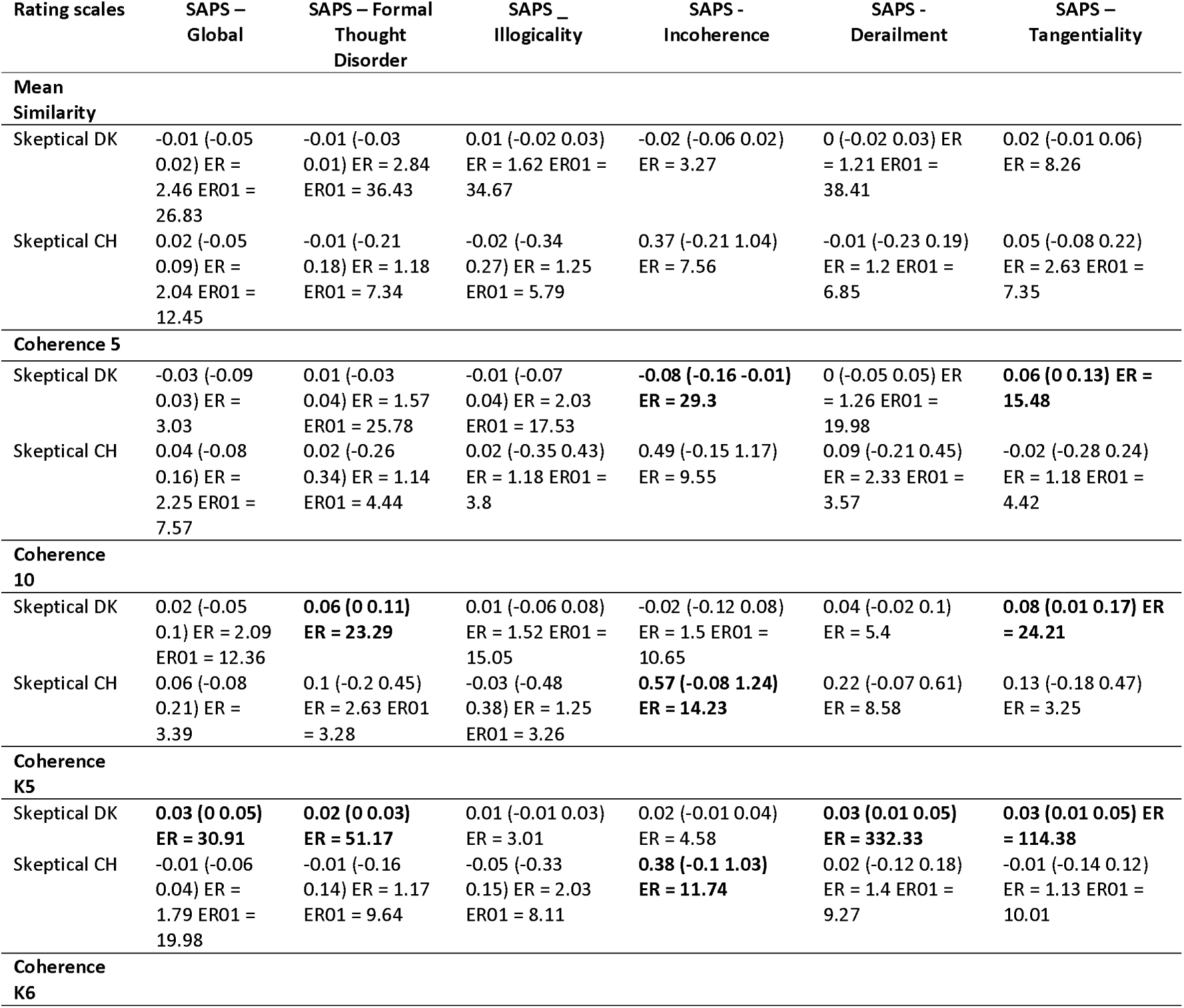
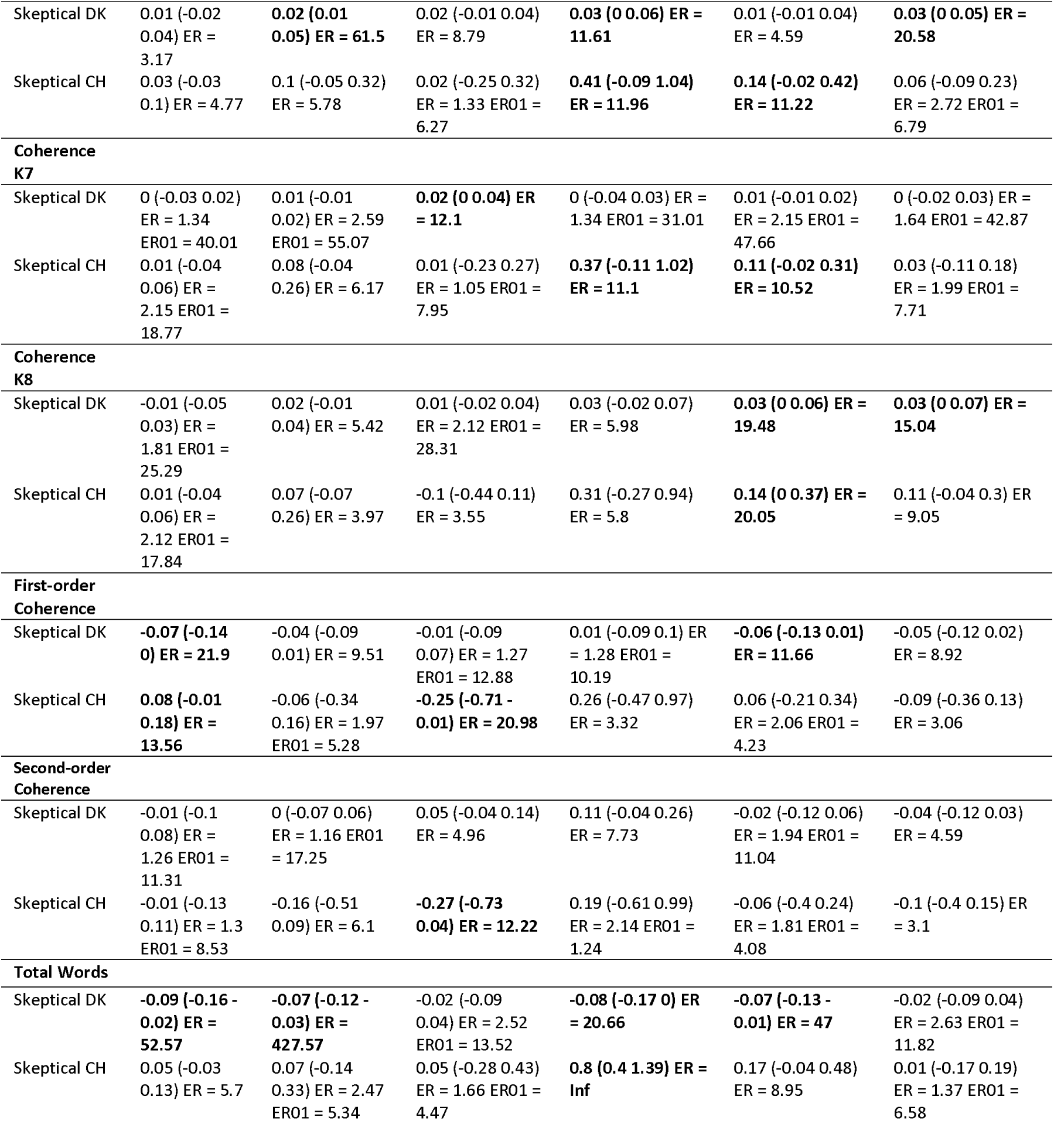
Estimated standardized relation between coherence measures and clinical features (SAPS). ER indicates the evidence ratio for the difference, ER01 the evidence ratio for the null effect.

**Table 5.** Estimated standardized relation between coherence measures and clinical features (SANS). ER indicates the evidence ratio for the difference, ER01 the evidence ratio for the null effect.

| Rating scales | SANS - Global | SANS - Alogia | SANS - Poverty of content | SANS - Poverty of speech |
| --- | --- | --- | --- | --- |
| **Mean Similarity** |  |  |  |  |
| Skeptical DK | -0.02 (-0.06 0.02) ER = 3.31 | -0.02 (-0.06 0.01) ER = 7.85 | -0.02 (-0.04 0.01) ER = 5.04 | **-0.04 (-0.09 -0.01) ER = 39.27** |
| Skeptical CH | -0.07 (-0.2 0.04) ER = 6.08 | -0.06 (-0.25 0.1) ER = 2.91 ER01 = 6.88 | 0.11 (-0.54 0.78) ER = 1.87 ER01 = 2.28 | -0.09 (-0.29 0.05) ER = 5.75 |
| **Coherence 5** |  |  |  |  |
| Skeptical DK | -0.02 (-0.09 0.05) ER = 1.92 ER01 = 13.29 | -0.02 (-0.07 0.03) ER = 2.43 ER01 = 16.81 | 0.02 (-0.03 0.07) ER = 3.11 | -0.04 (-0.11 0.02) ER = 6.33 |
| Skeptical CH | -0.12 (-0.32 0.06) ER = 6.41 | **-0.19 (-0.55 0.04) ER = 10.88** | 0.23 (-0.45 0.92) ER = 3.13 | **-0.26 (-0.58 -0.06) ER = 63.52** |
| **Coherence 10** |  |  |  |  |
| Skeptical DK | 0.06 (-0.03 0.16) ER = 7.08 | 0.05 (-0.02 0.12) ER = 6.78 | **0.07 (0.01 0.13) ER = 27.04** | 0.03 (-0.05 0.13) ER = 2.52 ER01 = 10.45 |
| Skeptical CH | -0.08 (-0.31 0.15) ER = 2.51 ER01 = 4.06 | -0.14 (-0.48 0.14) ER = 4.14 | 0.27 (-0.43 0.96) ER = 3.61 | -0.26 (-0.63 -0.01) ER = 24.32 |
| **Coherence K5** |  |  |  |  |
| Skeptical DK | 0.04 (0.02 0.07) ER = 299 | 0.03 (0.01 0.05) ER = 87.24 | 0.03 (0.01 0.04) ER = 135.36 | 0.01 (-0.01 0.05) ER = 3.61 |
| Skeptical CH | -0.08 (-0.18 0) ER = 21.22 | -0.09 (-0.27 0.04) ER = 6.98 | 0.36 (0 0.95) ER = 19.55 | -0.12 (-0.32 -0.01) ER = 31.79 |
| **Coherence K6** |  |  |  |  |
| Skeptical DK | 0.04 (0 0.07) ER = 25.32 | 0.02 (-0.01 0.05) ER = 8.45 | 0.01 (-0.01 0.04) ER = 6.24 | 0.02 (-0.01 0.07) ER = 7.23 |
| Skeptical CH | -0.03 (-0.15 0.07) ER = 2.49 ER01 = 8.6 | -0.05 (-0.25 0.1) ER = 2.57 ER01 = 6.95 | 0.29 (-0.23 0.9) ER = 6.72 | -0.13 (-0.36 0) ER = 17.46 |
| **Coherence K7** |  |  |  |  |
| Skeptical DK | 0.02 (-0.01 0.05) ER = 8.98 | 0.01 (-0.02 0.03) ER = 2.03 ER01 = 43.71 | 0 (-0.02 0.02) ER = 1.73 ER01 = 47.77 | 0 (-0.03 0.02) ER = 1.44 ER01 = 42.9 |
| Skeptical CH | -0.03 (-0.11 0.05) ER = 2.44 ER01 = 11.22 | -0.01 (-0.16 0.12) ER = 1.17 ER01 = 10.28 | 0.1 (-0.51 0.72) ER = 1.91 ER01 = 2.42 | -0.04 (-0.21 0.09) ER = 2.55 ER01 = 8.76 |
| **Coherence K8** |  |  |  |  |
| Skeptical DK | 0.03 (-0.02 0.08) ER = 6.04 | 0.04 (0.01 0.09) ER = 38.47 | 0.03 (0 0.06) ER = 16 | 0.03 (-0.02 0.09) ER = 4.67 |
| Skeptical CH | -0.02 (-0.11 0.06) ER = 1.97 ER01 = 11.02 | 0.01 (-0.15 0.18) ER = 1.31 ER01 = 8.48 | 0.05 (-0.61 0.73) ER = 1.41 ER01 = 2.68 | 0 (-0.16 0.15) ER = 1.08 ER01 = 9.78 |
| **First-order Coherence** |  |  |  |  |
| Skeptical DK | -0.15 (-0.26 -0.05) ER = 239 | -0.09 (-0.17 -0.02) ER = 89.91 | -0.07 (-0.13 -0.01) ER = 31.09 | -0.07 (-0.18 0.01) ER = 11.15 |
| Skeptical CH | -0.23 (-0.41 -0.09) ER = 170.43 | -0.3 (-0.64 -0.09) ER = 161.16 | -0.16 (-0.85 0.56) ER = 2.27 ER01 = 1.71 | -0.28 (-0.63 -0.05) ER = 50.72 |
| **Second-order Coherence** |  |  |  |  |
| Skeptical DK | -0.1 (-0.21 0) ER = 16.49 | -0.08 (-0.16 -0.01) ER = 43.44 | -0.05 (-0.13 0.02) ER = 5.76 | -0.05 (-0.13 0.03) ER = 6.96 |
| Skeptical CH | -0.33 (-0.57 -0.13) ER = 314.79 | -0.23 (-0.54 -0.02) ER = 27.71 | -0.01 (-0.72 0.73) ER = 1.04 ER01 = 1.72 | -0.27 (-0.58 -0.06) ER = 85.96 |
| **Total Words** |  |  |  |  |
| Skeptical DK | -0.1 (-0.18 -0.02) ER = 36.04 | -0.1 (-0.18 -0.04) ER = 351.94 | -0.04 (-0.1 0.02) ER = 7.2 | -0.09 (-0.18 -0.02) ER = 49.42 |
| Skeptical CH | -0.1 (-0.24 0.02) ER = 9.85 | -0.17 (-0.47 0.02) ER = 12.86 | 0.45 (0.03 1.08) ER = 23.79 | -0.24 (-0.54 -0.07) ER = 192.55 |

## Discussion

Current developments in clinical Natural Language Processing promise to revolutionize clinical practice. However, the scientific and clinical impact of these developments rely on the possibility to generalize NLP-based results across different samples and contexts. In this study, we assessed how the results of previous NLP studies measuring semantic coherence in schizophrenia generalize to a large novel cross-linguistic corpus. Globally, we found that only one previous result, i.e., reduced second-order coherence in schizophrenia, generalized across our entire corpus. Other results were replicated only for some specific languages: Chinese and German patients with schizophrenia showed lower coherence than controls across several, but not completely overlapping, measures; while Danish patients showed a mixed pattern with higher semantic coherence than controls across multiple measures, and lower coherence in first- and second-order coherence.

A first possible explanation is *clinical heterogeneity*. Schizophrenia is a highly heterogeneous disorder (Gratton & Mittal, 2020; Hitczenko et al., 2021; Schnack, 2019), and participants from different studies are likely to have different clinical profiles, thus contributing to inconsistent results. In particular, reduced semantic coherence has traditionally been associated with formal thought disorders and positive symptoms more in general (e.g. Andreasen, 1979). However, this assumption has only inconsistently been supported by the literature, with highly varying statistical significance and size of the effects found across studies: while some studies found a strong correlation between semantic coherence and measures of formal thought disorder (Bilgrami et al., 2022; Elvevåg et al., 2007), most others reported uncertain results (Bedi et al., 2015; Haas et al., 2020; Just et al., 2020; Morgan et al., 2021; Pauselli et al., 2018; Sarzynska-Wawer et al., 2021; Tang et al., 2021). Our study emphasized the lack of a clear picture. We found indeed only inconsistent associations between semantic coherence and formal thought disorders. Surprisingly, we found more reliable associations of coherence with higher ratings of negative symptoms (alogia, poverty of content). These results, overall, may indicate that the relationship between NLP measures and clinical ratings of symptoms is affected by several factors, such as the rating scale used, the clinical characteristics of the specific sample, the NLP metrics employed, and the statistical design adopted (e.g., ordinal modeling vs. group splitting).

A second possible explanation is *socio-demographic heterogeneity*. Gender, age and socio-economic status are known to impact speech production and therefore likely semantic coherence, and relatedly to affect the expression of specific symptoms such as thought disorder (e.g., Palaniyappan, 2021). Indeed, recent studies seem to suggest that NLP algorithms, and coherence measures in particular, can be biased by socio-demographic variables such as racial identity. For instance, Hitczenko et al. (2022) found the NLP algorithms rated entirely asymptomatic Black American participants as having language patterns consistent with thought disorder, thus leading to biased predictions. Because not all previous studies systematically adjusted their estimates for all key socio-demographic variables, we could only speculate that they affected previous findings. However, our results show that gender, age and education do affect the difference by group in coherence patterns.

A third possible explanation is *cross-linguistic (and relatedly cultural) variation*. Different languages present different linguistic structures and usage patterns (Evans & Levinson, 2009), and indeed computational measures of different linguistic aspects, including semantic coherence, have been shown to vary across languages (Palaniyappan, 2021; Sumiyoshi et al., 2004, 2014; Wydell & Butterworth, 1999; Dideriksen, et al., 2020). Nevertheless, previous literature has implicitly assumed the existence of NLP markers of schizophrenia independent of language and cultural groups analyzed. Our findings disconfirm this preconception. For example, Danish patients were generally more coherent than controls, whereas Chinese and German patients were less coherent than controls. One could speculate that this is driven by the general opacity of the Danish sound structure, which has been shown to relate to higher redundancy in speech (Dideriksen et al., 2020; Trecca et al., 2021). However, we still lack an overview across diverse languages and even more a systematic theory-driven approach that would identify relevant cross-linguistic contrasts to assess the impact of linguistic constraints on semantic coherence and other linguistic measures (e.g.,Çokal et al., 2019; Deffner et al., 2021).

Finally, the *variety in measures employed* in assessing clinical features as well as semantic coherence could be contributing to inconsistent results. Different clinical scales do not completely overlap in terms of included symptoms and symptom definition, and likely imply different representations of the underlying psychopathological dimensions (e.g., Marder & Galderisi, 2017). For example, Bedi et al. (2015) have found a significant association only between a specific combination of symptoms and linguistic measures, including coherence: this may suggest as measures of coherence may be related to more complex psychopathological dimensions than to specific symptoms, and they can vary across scales. Not least, we found important differences between the different NLP measures of coherence, both within and between languages. For example, while we found that Danish patients showed lower coherence than controls on first- and second-order coherence, they instead showed higher coherence on the different coherence-k measures. Coherence-k represents semantic similarity between single words irrespective of the sentence structure, while first- and second-order coherence represents semantic similarity between different sentences. Our results show how measuring semantic coherence at different levels of granularity can yield very different results and highlight the importance of using different NLP measures when assessing patterns of coherence in schizophrenia. They also point to the need for validation studies (e.g., Bilgrami et al., 2022) specifically aimed at assessing the psychometric properties of the different coherence measures, their relationship to clinical ratings, and their relationship to each other. Indeed, our robustness analysis have shown as different preprocessing options (e.g. transcript length and punctuation) can affect the various coherence measures. This is in line with previous previous studies (Elvevåg et al., 2007; Iter et al., 2018) and with recent evidence showing how some of these features (sentence length) can interact with socio-demographic characteristics and generate bias (Hitczenko et al., 2022).

We thus advocate for larger theory-driven validation studies incorporating socio-demographic, linguistic and clinical variability, in order to consistently identify and statistically account for sources of variation in the decreased semantic coherence. A promising venue is the establishing of large normative datasets in order to determine whether deviations from normative values have clinical significance, or they only reflect the characteristics of a specific sample (Marquand et al., 2016, 2019).

### Self-correcting approach

In this study we provided a concrete application of a cumulative yet self-critical scientific approach. We relied on a review of previous literature to design the current study and to set up priors for our analyses. At the same time, we critically attempt to replicate previous findings and compare statistical inferences relying on informed priors with inferences relying on skeptical priors. Our findings highlight some of the issues with previous literature. Indeed, we found only partial replications and informed models were often less robust and generalizable to new data than skeptical ones. Both pieces of evidence point towards a lack of generalizability of the previous literature, be it due to unreliable estimates, sparsity of previous data or lack of representativity of the previous samples compared to the current ones. Thus, even though the informed priors per se are not directly useful to generally improve our estimates, the comparison with skeptical ones provides valuable checks of the inferential robustness and of how the current study relates to the previous literature.

### Limitation and future perspectives

One of the limitations of the present study is that we used pre-trained algorithms to quantify semantic coherence. This may have limited the sensitivity of the algorithms in detecting linguistic patterns specific to the characteristics of our corpus. On the other hand, using pre-trained algorithms allowed us to compare different coherence measures across different samples and languages in a scalable and easily replicable way. Another limitation is that we focused on and compared single measures of semantic coherence: future studies should focus more on developing replicable (i.e., freely accessible and computationally feasible) machine learning pipelines to test the generalizability of large multidimensional patterns of features to account for shared variance across features, speech tasks, and languages. Finally, another limitation is the large variability in terms of clinical and demographic features of our multilingual corpus. While this variability may have contributed to the differences in the main results, we argue that the sample size of the samples of our corpora is larger than average (median for participants with schizpohrenia = 34.5), and that this setting provided us with a concrete basis for evaluating the heterogeneity of NLP measures across samples and languages.

## Conclusions

In this study we showcased how a cumulative, self-correcting and replicable approach can be used to test the generalizability and robustness of NLP results in schizophrenia across different languages, samples and measures. Overall, we found large cross-linguistic variability in NLP-based assessments of semantic coherence in schizophrenia, with different sources of heterogeneity interacting at different levels. Future studies should take this variability into account in order to devise effective clinical applications able to target different ranges of patients and identify the presence of potential bias.

## Data Availability

The original speech transcripts cannot be shared as they are considered identifiable data, in line with our consent forms and current data privacy regulations.

https://osf.io/8btp6/

