## Supplementary Material for "Speech disturbances in schizophrenia: assessing cross-linguistic generalizability of NLP automated measures of coherence"

<u>The supplementary materials contain the following sections:</u>

- SM1: Description of the speech corpus
- SM2: Animated Triangles Task (ATT)
- SM3: Speech preprocessing and data preparation
- SM4: Systematic review of the literature and NLP quantification of coherence measures
- SM5 Summary effects sizes, model priors, model fitting and model assessment, IQR results, association between coherence measures and symptoms
- SM6: Robustness analysis
- SM7: Software implementation notes

**SM1 – Description of the speech corpus**

**Danish corpus**

The patient and control samples for the present study were collected within three consecutive studies recruiting patients at the same clinical location, i.e., at OPUS - Clinic for people with schizophrenia, Aarhus University Hospital Risskov, in the period from 2009 to 2018 (Beck et al., 2020; Bliksted et al., 2017; Veddum et al., 2019).

The patients had been diagnosed according to ICD-10 criteria by experienced psychiatrists. Exclusion criteria were a history of neurological disorder, severe head trauma, or substance abuse problem according to the ICD-10. Patients were excluded if they did not understand spoken Danish sufficiently to understand the testing procedures or if they had an estimated premorbid IQ below 70 based on their history.

All patients with schizophrenia, except the sample reported in Veddum et al. (2019)[3], were interviewed with the Scale for Assessment of Negative Symptoms (SANS), the Scale for Assessment of Positive Symptoms (SAPS), and the Personal Social Performance Scale (PSP, a measure of social functioning in schizophrenia).

**IQ**

Verbal intelligence was estimated from two subtests (Vocabulary and Similarities) from WAIS-III (Wechsler Adult Intelligence Scale, Third edition Wechsler, 1997). The two subtests were chosen based on their high correlation with the verbal WAIS-III IQ-score.

**Control groups**

Patients and non-clinical controls were matched by age, sex, and level of education and parental socioeconomic status (except sample 3, matched only by age and sex). Control participants did not have a history of neurological or mental illness (and neither had their first-degree relatives), severe head injury or drug- or alcohol dependence according to the ICD-10 criteria. Demographics and social functioning of the patients and their matched controls are summarized in **Table 1**.

**Ethics**

The participants received written and oral information about the project, and written informed consent was obtained before inclusion. The study was approved by The Central Denmark Region Committees on Biomedical Research Ethics (Ref: M-2009–0035; Ref: 2007-58-0010) and the Danish Data Protection Agency. The project complied with the Helsinki Declaration of 1975, as revised in 2008.

**Chinese corpus**

The patient and control samples were collected within two consecutive studies recruiting patients at the same clinical location, i.e., Renmin Hospital of Wuhan University (Beck et al., 2020; Yang et al., 2017), and a part of the data (n = 41 participants) has not yet been published.

Patients met the diagnostic criteria for schizophrenia according to the ICD-10 and were diagnosed by psychiatrists.

Exclusion criteria were a history of severe head trauma or neurological illness or if they had a substance abuse problem according to the ICD-10. Patients with an estimated premorbid IQ below 70 based on previous history or who were unable to understand spoken Chinese well enough to understand testing instructions were also excluded. All of the Chinese patients were of Han Chinese ethnicity.

Patients were interviewed with the Scale for Assessment of Negative Symptoms (SANS), the Scale for Assessment of Positive Symptoms (SAPS) and the Personal Social Performance Scale (PSP).

**IQ**

Verbal intelligence was estimated from Vocabulary subtests from WAIS-III (Wechsler Adult Intelligence Scale, Third edition) (Wechsler, 1997). The subtest was chosen based on its high correlation with the verbal WAIS-III IQ-score: (Wechsler, 1997).

**Control group**

There were no differences between patients and non-clinical controls regarding age and sex at group level. The exclusion criteria for the healthy subjects were the same as for patients. In addition, healthy control subjects were excluded if they, or a first-degree relative, met any psychiatric diagnosis according to the ICD-10.

**Ethics**

All participants received written and spoken information about the project, and written consent was obtained. The project was approved by the Ethics committee of Renmin Hospital of Wuhan University and the Institutional Review Board of the Institute of Psychology, the Chinese Academy of Sciences. The authors assert that all procedures contributing to this work comply with the ethical standards of the relevant national and institutional committees on human experimentation and with the Helsinki Declaration of 1975, as revised in 2008.

**German corpus**

The patient and control samples for the present study were collected in a study (studies recruiting patients at the University of Muenster and the Psychiatric Hospital of the Regional Association of Westphalia-Lippe (Koelkebeck et al., 2010) (LWL).

The patients had been diagnosed with schizophrenia by experienced psychiatrists using the Structured Clinical Interview for DSM-IV (SCID-I). Psychopathology was assessed with the Positive and Negative Syndrome Scale (PANSS).

Patients with any history of other psychiatric disorders, neurological disorders, serious head injury, alcohol or illegal drug abuse, or insufficient knowledge of the German language were excluded from the study.

**IQ**

Verbal intelligence was estimated from Vocabulary subtests from WAIS-III (Wechsler Adult Intelligence Scale, Third edition) (Wechsler, 1997). The subtest was chosen based on its high correlation with the verbal WAIS-III IQ-score: (Wechsler, 1997).

**Control group**

Healthy controls were matched with patients for age, sex and education. Healthy participants with no history of Axis I DSM-IV diagnoses (SCID-I), illegal drug use, alcohol abuse or addiction, or neurological disorders served as a control group. Subjects with any first-degree relatives with a history of mental disorders were excluded from the study.

**Ethics**: After hearing a complete description of the study, written informed consent was obtained from all participants. The study was approved by the Ethics Committee of the University of Muenster and the State Chamber of Physicians of Nordrhein-Westphalia and has been carried out in accordance with the Declaration of Helsinki.

**SM2 – Animated Triangles Task**

Voice recordings were collected using the Animated Triangles Task (Abell et al., 2000; Bliksted et al., 2016; Castelli et al., 2000). The task is generally used to measure theory of mind (ToM) and involves video clips representing an interaction between animated geometrical shapes (triangles). In some of the clips the two triangles are moving randomly and unintentionally (e.g., bouncing off walls) (4 clips). In the other clips, the triangles are interacting intentionally to influence the mental state of one another (e.g., a larger triangle trying to convince a small one to leave a closure) (4 clips) or merely performing an activity alone or together (4 clips). The duration of each animation is approximately 40 seconds. The participants were asked to provide an interpretation of what was going on in each animation and their answers were audio-recorded and then transcribed. Not all studies used the 4 pure Action clips.

**SM3 – Speech preprocessing**

The answers of participant and interventions of the examiner were audio-recorded, and then all audio recordings were manually time coded, to identify the segments corresponding to the participants' video description, thus excluding instructions, prompts (e.g., "Can you say anything more about that?") as well as questions, backchannels (such as 'hmmm', or 'ok', etc.) provided by the examiner. The audio-recordings were then transcribed by research assistants. After the transcription, interventions from the examiner were removed, as well as punctuations and fillers such as 'uhm' and ”ehm". No other preprocessing was performed; specifically, repetitions were not removed.

The data were then prepared for computer-based analyses by using custom R scripts relying on the UDPipe Natural Language Processing - Text Annotation in R (Straka et al., 2016). UDPipe is an opensource and trainable pipeline parsing system. It performs sentence segmentation, tokenization, part-of-speech tagging, lemmatization, morphological analysis, and dependency parsing

We then used fastText pre-trained models for the different languages (Bojanowski et al., 2017) to vectorize the tokenized speech samples, yielding a 300-dimensional vector for each word. After that, we computed semantic coherence between words, i.e., word-to-word similarity, by calculating the cosine similarity between the corresponding vectors associated with each word. The cosine similarity values range between − 1 and 1, with − 1 representing the lowest similarity and 1 the highest similarity between two words.

**Table SM3_A.** Frequency of words, mean sentence length, and mean number of sentences for patients with schizophrenia (SCZ) and controls (HC) in the different languages (Danish, German, and Chinese).

| Feature | Danish | | German | | Chinese | |
|---|---|---|---|---|---|---|
| | SCZ | HC | SCZ | HC | SCZ | HC |
| Total number of words (per trial) | 41.03 (35.3) | 49.2 (37.1) | 36.73 (24.9) | 39.51 (25.5) | 39.28 (38.7) | 51.14 (41.2) |
| Mean sentence length (in words) | 17.10 (16.1) | 19.71 (16.8) | 16.76 (13.0) | 25.65 (18.8) | 11.68 (8.1) | 12.4 (7.8) |
| Mean number of sentence (per trial) | 2.40 (2.0) | 2.50 (1.7) | 1.54 (0.9) | 2.19 (1.28) | 4.13 (2.92) | 3.36 (2.72) |

**Table SM3_B**. Total and relative frequency of the unique parts of speech (UPOS), for language and diagnosis, before data cleaning.

| UPOS | Language | Total frequency (UPOS) | Total frequency | Relative frequency | Total frequency (UPOS) | Total frequency | Relative frequency |
|---|---|---|---|---|---|---|---|
| Diagnosis | | Controls | | | Patients with schizophrenia | | |
| ADJ | CH | 503 | 19963 | 0.0251966137 | 408 | 19629 | 0.0207855724 |
| ADJ | DK | 4164 | 63681 | 0.0653884204 | 3035 | 45654 | 0.0664782932 |
| ADJ | GE | 1228 | 1433 | 0.0856644576 | 1044 | 13351 | 0.0781963898 |
| ADP | CH | 895 | 19963 | 0.0448329409 | 838 | 19629 | 0.0426919354 |
| ADP | DK | 5073 | 63681 | 0.0796626937 | 3336 | 45654 | 0.0730713629 |
| ADP | GE | 906 | 14335 | 0.0632019533 | 576 | 13351 | 0.0431428357 |
| ADV | CH | 1936 | 19963 | 0.0969794119 | 1415 | 19629 | 0.0720872179 |
| ADV | DK | 11026 | 63681 | 0.1731442660 | 8104 | 45654 | 0.1775090901 |
| ADV | GE | 2202 | 14335 | 0.1536100453 | 1547 | 13351 | 0.1158714703 |
| AUX | CH | 542 | 19963 | 0.0271502279 | 474 | 19629 | 0.0241479444 |
| AUX | DK | 2428 | 63681 | 0.0381275420 | 1729 | 45654 | 0.0378718185 |
| AUX | GE | 1145 | 14335 | 0.0798744332 | 1057 | 13351 | 0.0791700996 |
| CCONJ | CH | 57 | 19963 | 0.0028552823 | 70 | 19629 | 0.0035661521 |
| CCONJ | DK | 4013 | 63681 | 0.0630172265 | 2797 | 45654 | 0.0612651684 |
| CCONJ | GE | 729 | 14335 | 0.0508545518 | 583 | 13351 | 0.0436671410 |
| DET | CH | 706 | 19963 | 0.0353654260 | 802 | 19629 | 0.0408579143 |
| DET | DK | 4919 | 63681 | 0.0772443900 | 3621 | 45654 | 0.0793139703 |
| DET | GE | 1690 | 14335 | 0.1178932682 | 1285 | 13351 | 0.0962474721 |
| FILL | CH | 418 | 19963 | 0.0209387367 | 274 | 19629 | 0.0139589383 |
| FILL | DK | 1303 | 63681 | 0.0204613621 | 1034 | 45654 | 0.0226486179 |
| FILL | GE | 1312 | 14335 | 0.0915242414 | 1700 | 13351 | 0.1273312860 |

| | | | | | | | |
|---|---|---|---|---|---|---|---|
| INTJ | DK | 33 | 63681 | 0.0005182079 | 28 | 45654 | 0.0006133088 |
| NOUN | CH | 4119 | 19963 | 0.2063317137 | 4350 | 19629 | 0.2216108819 |
| NOUN | DK | 6207 | 63681 | 0.0974702030 | 4165 | 45654 | 0.0912296841 |
| NOUN | GE | 1252 | 14335 | 0.0873386815 | 910 | 13351 | 0.0681596884 |
| NUM | CH | 1163 | 19963 | 0.0582577769 | 1435 | 19629 | 0.0731061185 |
| NUM | DK | 460 | 63681 | 0.0072235047 | 292 | 45654 | 0.0063959346 |
| NUM | GE | 19 | 14335 | 0.0013254273 | 5 | 13351 | 0.0003745038 |
| PART | CH | 2165 | 19963 | 0.1084506337 | 2358 | 19629 | 0.1201283815 |
| PART | DK | 710 | 63681 | 0.0111493224 | 457 | 45654 | 0.0100100758 |
| PART | GE | 310 | 14335 | 0.0216253924 | 265 | 13351 | 0.0198487005 |
| PRON | CH | 914 | 19963 | 0.0457847017 | 808 | 19629 | 0.0411635845 |
| PRON | DK | 7205 | 63681 | 0.1131420675 | 5328 | 45654 | 0.1167039033 |
| PRON | GE | 1091 | 14335 | 0.0761074294 | 992 | 13351 | 0.0743015504 |
| PROPN | CH | 98 | 19963 | 0.0049090818 | 84 | 19629 | 0.0042793825 |
| PROPN | DK | 20 | 63681 | 0.0003140654 | 15 | 45654 | 0.0003285583 |
| PROPN | GE | 102 | 14335 | 0.0071154517 | 65 | 13351 | 0.0048685492 |
| PUNCT | CH | 3179 | 19963 | 0.1592446025 | 3407 | 19629 | 0.1735697183 |
| PUNCT | DK | 7665 | 63681 | 0.1203655721 | 5822 | 45654 | 0.1275244228 |
| PUNCT | GE | 358 | 14335 | 0.0249738403 | 1687 | 13351 | 0.1263575762 |
| SCONJ | DK | 1340 | 63681 | 0.0210423831 | 827 | 45654 | 0.0181145135 |
| SCONJ | GE | 249 | 14335 | 0.0173700732 | 205 | 13351 | 0.0153546551 |
| VERB | CH | 3263 | 19963 | 0.1634523869 | 2902 | 19629 | 0.1478424780 |
| VERB | DK | 7043 | 63681 | 0.1105981376 | 5010 | 45654 | 0.1097384676 |
| VERB | GE | 1619 | 14335 | 0.1129403558 | 1322 | 13351 | 0.0990188001 |

**Note:** ADJ: adjective, ADP: adposition ADV: adverb AUX: auxiliary CCONJ: coordinating conjunction DET: determiner INTJ: interjection NOUN: noun NUM: numeral PART: particle PRON: pronoun PROPN: proper noun PUNCT: punctuation SCONJ: subordinating conjunction SYM: symbol VERB: verb**;** Danish (DK), German (GE) and Chinese (CH).

### SM4 – Systematic review of the literature and NLP quantification of coherence measures

We systematically screened the current literature relying on - but not fully complying with[1] - the indications of the Preferred Reporting Items for Systematic Reviews and Meta-Analyses Guidelines (PRISMA) for transparent reporting of a systematic review. We conducted a literature search on PubMed and Google Scholar by using the following search terms ((Automated language analysis) or (natural language processing) or (Automatic Data Processing) or (semantic coherence) or (discourse coherence) or (vector semantics) or (speech coherence) or (latent semantic)) and ((schizo*) or (psychosis)). The search was conducted on 19/05/2021 and updated on Google Scholar on 18/11/2021. We complemented the list by performing a backward and forward literature search: we screened the bibliography of the papers and recent reviews (Argolo et al., 2020; Corcoran et al., 2020; Corcoran & Cecchi, 2020; de Boer et al., 2018; De Boer et al., 2020; Hitczenko et al., 2021) found in the search, and the papers citing them as identified by Google Scholar. Articles were screened for eligibility by two authors (A.P and J.M.L). Study selection was conducted according to

[1] Here a list of differences compared to full compliance to the PRISMA checklist: a) we have not directly contacted the authors requesting relevant missing estimates; b) we have not provided an assessment of risk of bias for each study included; c) we have not analyzed the role of potential moderators, such as speech task, or socio-demographic features; d) we have not assessed publication bias and influential studies, due to the aim of our review. We aimed at providing a systematic synthesis of the previous literature, identifying the NLP measures of coherence used in previous studies, and extracting relevant summary estimates for those measures.

the following inclusion criteria: (a) empirical study, (b) automated measures, i.e., NLP-based, of coherence of participants with schizophrenia or schizoaffective disorder, or participants at high clinical risk of psychosis (HCR) (c) sample including at least two individuals with schizophrenia or schizoaffective disorder, (d) inclusion of a non-clinical comparison group. **Fig. SM4** shows the flow-diagram of study selection.

**Selection of coherence measures and comparison with previous studies**

Among the final set of studies, we selected scalable measures of coherence, i.e., easier to apply to a larger set of languages (without ad hoc wordlists, etc..), and to corpora with limited size (e.g., excluding training deep learning methods on the corpus). We found 14 studies using NLP measures to quantify semantic coherence in schizophrenia (see **Table SM4**). However, we were able to retrieve relevant estimates only from 5 studies including fourteen measures of semantic coherence (see **Fig. SM4**) which we used as informed priors in our analysis (see **SM5**). **Table SM4** reports a synthesis of results of previous studies (including also those not reporting relevant estimates) and qualitative comparisons with the results of the present research. Our implementation of the measures of semantic coherence sometimes deviates from the original studies, both in the data preprocessing, and in the word and sentence embeddings employed. For example, Corcoran et al., (2018) normalized the different coherence measures for sentence length, or different word embeddings have been employed in the different studies (for a detailed synthesis of the preprocessing methods and word and sentence embeddings employed in the different studies: https://osf.io/8btp6/). This choice and our selection of the coherence measures was motivated by the goal of the present research, that is to build a data analysis framework able to extract different measures of coherence and compare them in a scalable way across different languages. We opted to use median and interquartile range of semantic measures, contrary to more commonly used mean, standard deviation and range, because they are more robust to outliers and provide more complementary information in case of long tails in the distribution described. Median and interquartile range more robustly present independent measures of mode and variance of the distribution, respectively.

**Figure SM4.** Fig. SM4. Flow chart showing the literature search and study selection process following the indications of the Preferred Reporting Items for Systematic Reviews and Meta-analyses (PRISMA)

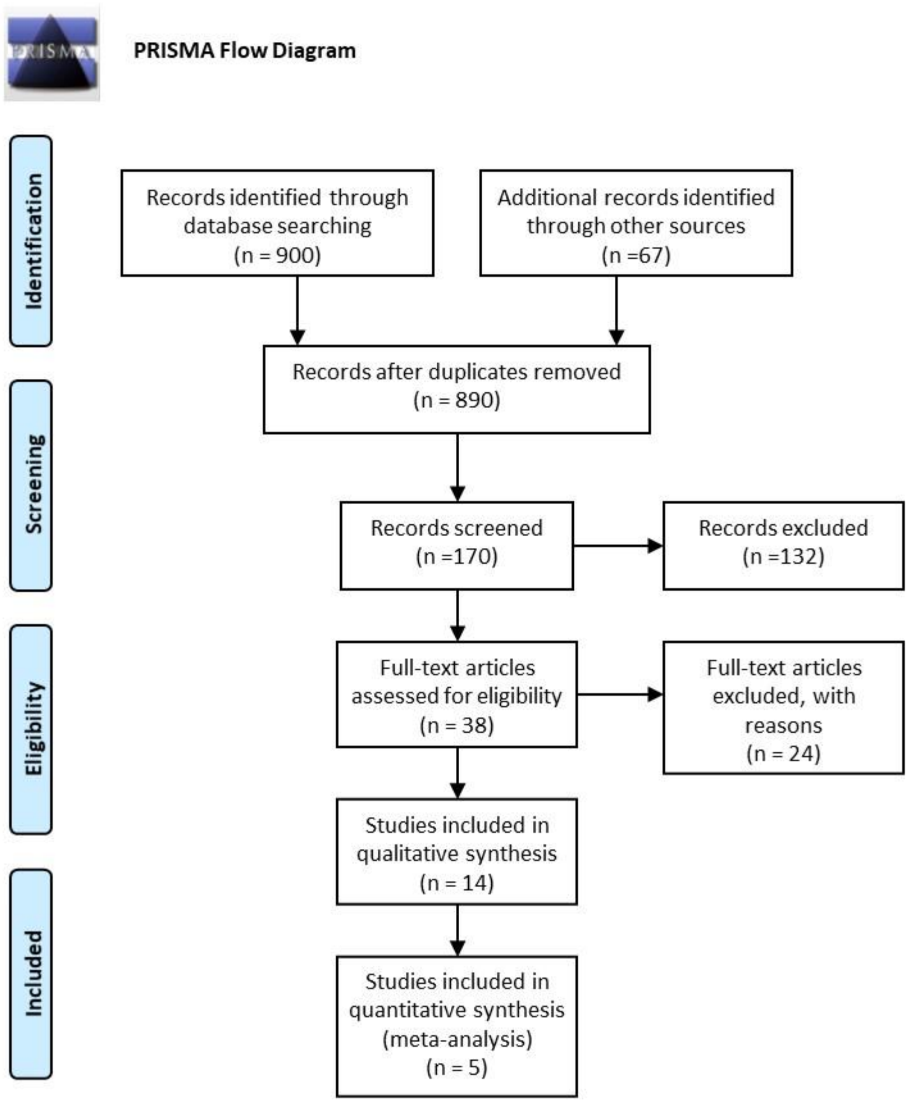

**Table SM4.** Definition of the fourteen NLP coherence measures used in previous studies in schizophrenia, references and results of the previous studies which employed these measures for comparing patients with schizophrenia (SCZ) and HC, and comparison with the results of the present study.

| NLP - Coherence Measures | Description | References | | |
|---|---|---|---|---|
| *Similarity Mean* | Average semantic similarity of each word to the immediately preceding word | Pauselli et al. (2018) | Lower coherence in SCZ | Lower coherence in CH, no difference in DK and GE |
| | | Khudyakova & Ryazanskaya, 2020 | NR | |
| | | Bar et al., 2019 | Lower coherence in SCZ | |
| *Coherence 5* | Average semantic similarity of each word in 5-words window | Pauselli et al. (2018) | No difference | Lower coherence in CH, no difference in DK and GE |
| *Coherence 10* | Average semantic similarity of each word in 10-words window | Pauselli et al. (2018) | No difference | Lower coherence in CH, higher coherence in DK, no difference in GE |
| *Coherence-K2*<br>*Coherence-K3*<br>*Coherence-K4*<br>*Coherence-K5*<br>*Coherence-K6*<br>*Coherence-K7*<br>*Coherence-K8*<br>*Coherence-k9*<br>*Coherence-k10* | Word-to-word variability at k inter-word distances | Bar et al. (2019) | Lower coherence in SCZ[2] | Lower coherence in CH and GE, higher coherence in DK, |
| | | Corcoran et al. (2018) | Lower coherence and higher variance coherence in SCZ | |
| | | Voppel et al., 2021 | Higher variance coherence in SCZ[3] | |
| *First-order Coherence* | Similarity of consecutive phrase vectors[3] | Bedi et al., (2015) | Lower coherence in SCZ | Lower coherence in CH and DK |
| | | Just et al., 2019 | No difference[4] | |
| | | Morgan et al. (2021) | Lower coherence in SCZ | |
| | | Haas et al. (2020) | No difference | |
| | | Iter et al. (2018) | No difference[4] | |
| | | Just et al. (2020) | Lower coherence in SCZ[5] | |

[2] The authors found lower coherence mean in schizophrenia for all the k-coherence measures including only content words, while including all words patients with SCZ showed lower coherence but not significantly different from controls

[3] Voppel et al. (2021) calculated coherence-k measures with a varying k-window size from 2 to 20. Since they found relevant differences between patients with SCZ and controls in the k-window interval comprised between 5 and 10 words, and since the aim of the present paper is to extract the coherence measures most easy to quantify, we limited here the selection of the k-windows interval between 2 and 10 words.

[4] The authors derived the coherence measures reported in Table SM4 using different sentence and words embeddings. We here report the results for sentence and words embeddings corresponding to those used in the present research, i.e., word2vec for word embeddings and mean vector for sentence embeddings.

[5] This result was found only for patient with higher level of positive formal thought disorder (FTD), while no difference is reported between patients with low level of FTD and controls

| | | | | |
|---|---|---|---|---|
| | | Sarzynska-Wawer et al., (2021) | NR | |
| *Second-order Coherence* | Similarity between phrases separated by another intervening phrase | Bedi et al. (2015) | Lower minimum coherence | Lower coherence in CH and DK |
| | | Sarzynska-Wawer et al., (2021) | NR | |

### SM5 – Summary effect sizes, <u>model priors, model fitting and analysis on symptoms</u>

#### <u>Summary effect sizes</u>

Summary effect sizes (ES) were calculated using the dataset available here: https://osf.io/8btp6/. For each coherence measure feature we extracted the estimate of the standardized mean difference (SMD; also known as Hedges' g) between individuals with schizophrenia and HC. The effects were then analyzed using 2-level hierarchical Bayesian regression models to estimate the summary effect sizes and corresponding credible (i.e., Bayesian confidence) intervals. We explicitly modeled the heterogeneity (or $\sigma^2$) in the results of the studies by varying effects by article and participants. A detailed description of the statistical procedure adopted is provided in Parola et al., 2020). This yielded summary effect sizes for: a) the effect of diagnosis for the following coherence measures (**Table SM4**). As for the coherence measures for which only a single study was available, we used the standardized mean difference (Hedges' g) between individuals with schizophrenia and HC as ES. We then used ES estimates as informed priors in the analysis. For ease of comparison, we standardized our coherence measures, that is, we centered them on the mean, and divided them by the standard deviation, separately for each language. Scaling within language has two important motivations. First, it makes our study more comparable with previous monolingual studies. Second, if one language were to present a different variance in a feature than other languages, cross-linguistic effects would be less comparable.

#### Analysis on differences between patients with schizophrenia and healthy controls

#### Model priors

We first specified the prior distributions for each parameter to be estimated. We adopt a weakly skeptical approach to priors, in that we aim at reducing the prior probability of extreme values for the parameters, while not overly influencing the inference.

Skeptical priors were specified as normal distribution centered at zero (no group difference), with a standard deviation of .5. The expected estimates were thus predominantly included within -.1.5 and 1.5, but could still be swayed by the data. Individual variability was modeled using a positive half-normal prior centered at 0, with a standard deviation of 0.7 from the estimate for the specific group and language of the individual, thus regularizing the inference. Sigma was modeled using a normal distribution centered at 0.3 with a standard deviation of 0.1. Informed priors - when available - were normal distributions based on the meta-analytic effects.

This choice of priors did stem from initial (potentially subjective) intuitions as to plausible ranges of difference in coherence measures between patients with schizophrenia and controls, based on previous experiences on similar modeling practices. However, these choices underwent extensive rigorous checks to ensure the priors to be adequate and only weakly informative (Gelman et al., 2020), that is, that the plausible values of coherence measures predicted by the model on the base of the priors only would not present odd biases. In particular, we first performed prior predictive checks, making predictions as to the data we would predict based on the priors and likelihood function only, before fitting the model to the actual data (see **Figure SM5_A**). After fitting the model to data (and before assessing the inferences in the model and testing hypotheses), we performed prior-posterior update checks. In other words, we plotted prior against posterior estimates for each parameter of interest and assessed the relative impact of the prior. This meant making sure that the posterior estimates had lower variance (more peaked and narrower) than the prior ones, and that they were well within the range covered by the prior expectations. In other words, we checked whether the model had learned from the data (posterior estimates being more confident than the priors), and whether the prior had unduly restrained the range of the posterior estimates (posterior estimates pushing against the low probability boundary set by the prior). For varying effects, which have more explicit regularizing functions (defining the amount of partial pooling across, in this case, participants within the same group), the second concern was more pressing than the former. Prior-posterior update checks are reported below (see **Figure SM5_B** and **Figure SM5_C**). Note that all priors were thus assessed and this motivated some adjustments. The adjustment of the priors is purely motivated by the need to ensure good model quality: an a priori reasonable region of the parameter space to search during the model fitting, the evaluation of potential biases introduced by priors in the inference, etc. The adjustment of priors is not motivated by the results, and it is indeed performed before any hypothesis testing.

**Figure SM5_A** –Plot of the prior predictive checks (light blue) and observed data (bold blue). Note that the three language groups (DK, GE, CH) are given the same prior expectations for all parameters, thus implicitly assuming no difference between them (so that any evidence for a difference that we might find would come exclusively from the data and not from the priors).

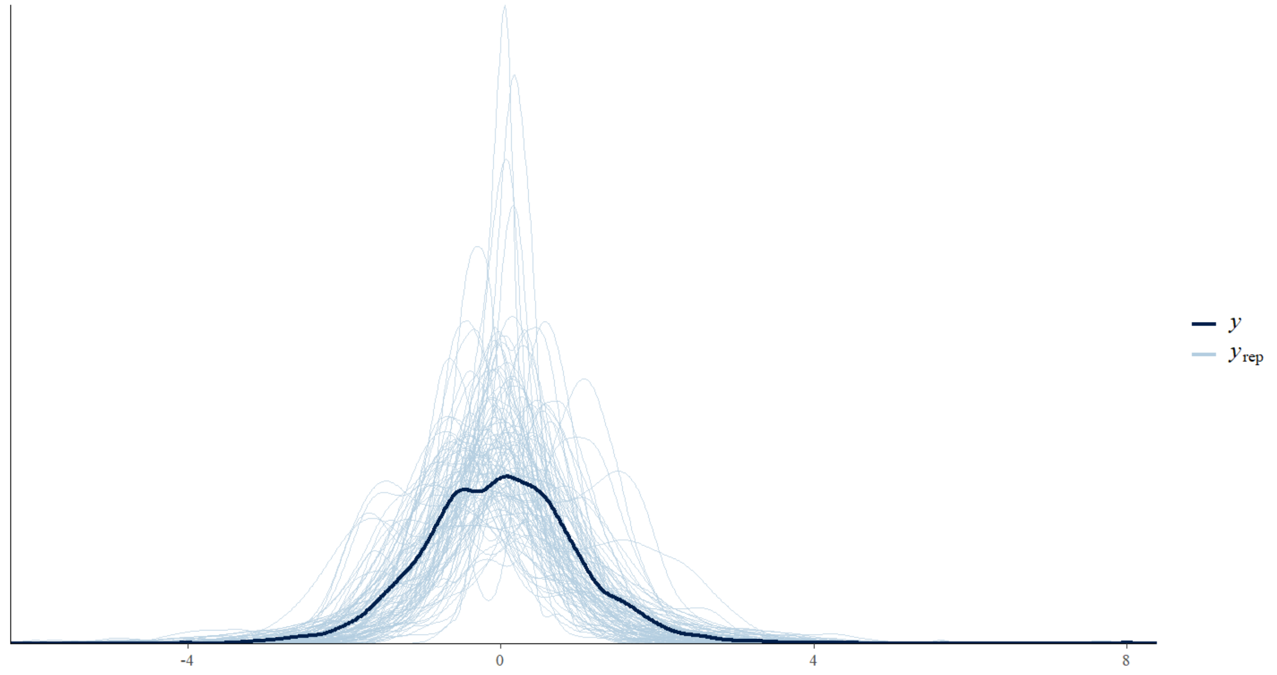

**FIGURE SM5_B.** Plot of the prior-posterior predictive check. Prior (light blue) and posterior (dark blue) distributions *for the intercept* in the model *for the Danish language*

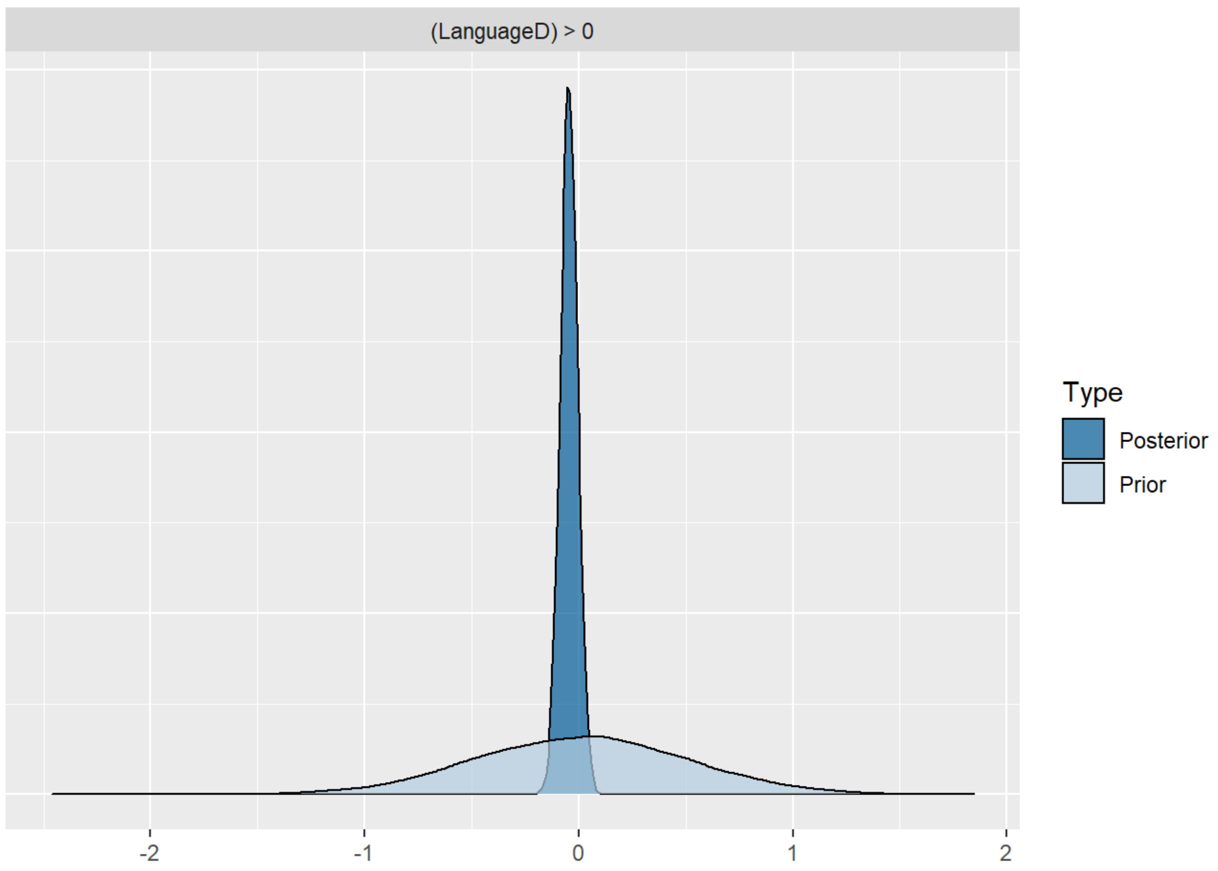

**FIGURE SM5_C.** Plot of the prior-posterior predictive check. Prior (light blue) and posterior (dark blue) distributions for the *effect of diagnosis (SCZ-HC) for the Danish language*

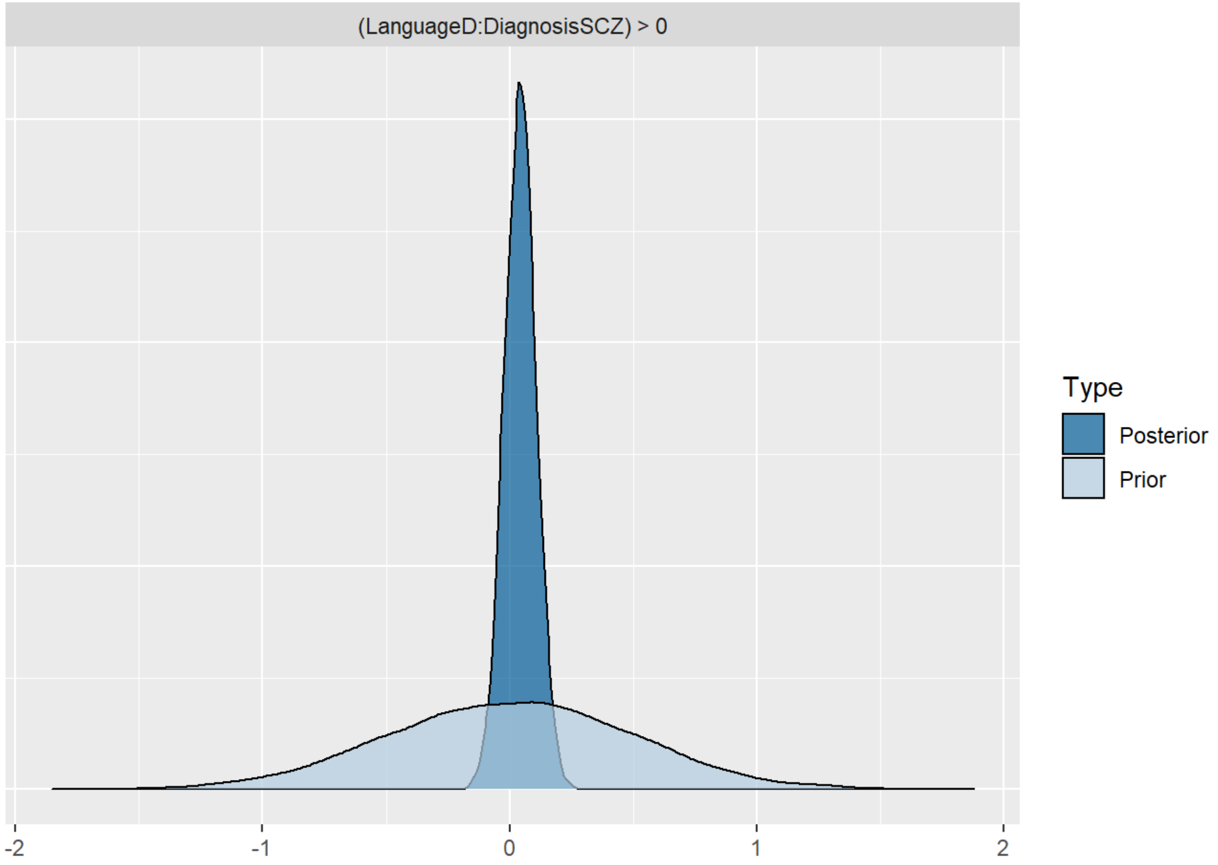


### Model fitting

The models were fitted using Hamiltonian MonteCarlo samplers with 2 parallel chains and 2,000 iterations each, an adapt delta of 0.99 and a maximum tree depth of 20 in order to ensure no divergence in the estimation process. The quality of the models was assessed beyond the prior predictive checks and prior-posterior update checks discussed in the previous paragraph. We performed posterior predictive checks (see **Figure SM5_D**), akin to traditional residual checks, to assess whether the model was able to capture the empirical distribution of the data and did not present obvious biases, such as overestimating low values. We made sure that R^ statistics were lower than 1.05, that is, that the independent Markov chains used to identify the posterior distributions converged on their estimates. We ensured that no divergences were generated in the process of estimation, that is, that the possible parameter values were satisfactorily explored, without specific ranges of values being excessively difficult to evaluate. We checked that the number of effective bulk and tail samples was above 200, that is, that the model fitting process was able to explore the possible parameter values and was able to sample at least 200 independent possible values. See figures below for predictive checks, and prior-posterior update checks.

**Figure SM5_D.** Plot of the posterior predictive check. We overlay the distribution of the actual data for the Coherence - 5 measure (bold blue line) with the distribution of 100 simulations from the fitted model (predictions, each simulation a pale blue line). This allows us to identify potential biases in the model.

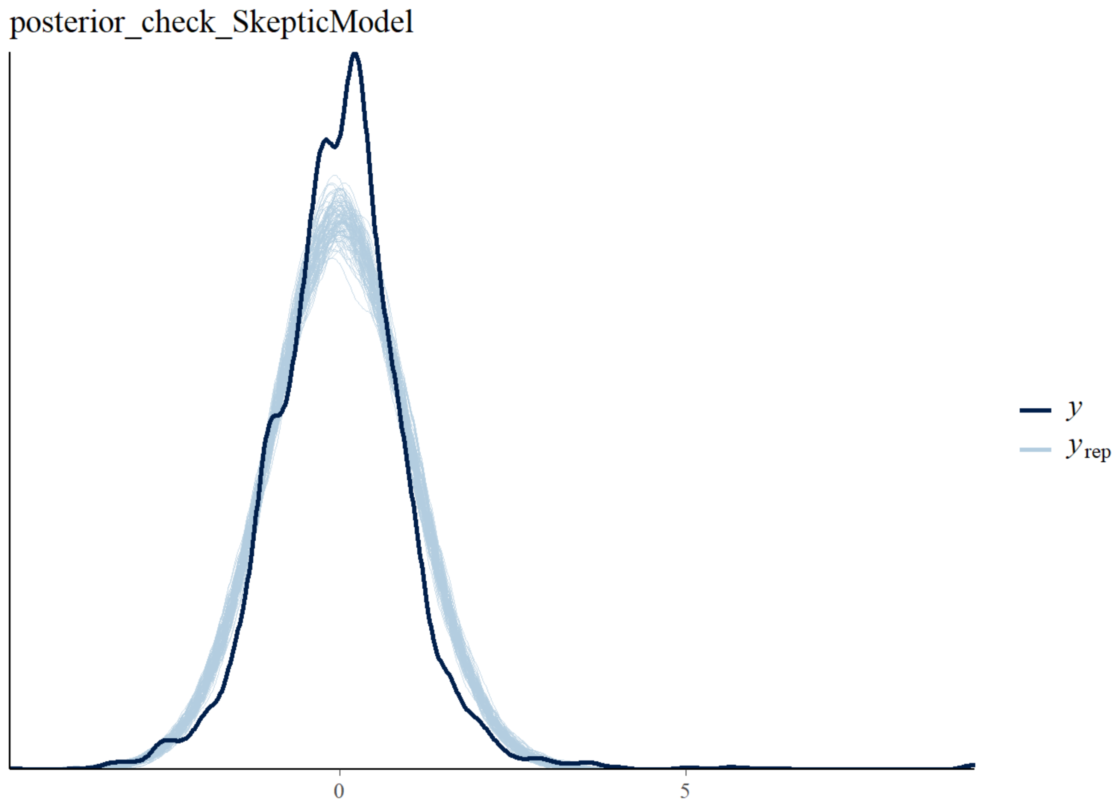


**Model assessment**

We then reported the estimated difference by group in terms of mean difference separately by language, 95% Compatibility Intervals (CIs, indicating the probable range of difference, assuming the model is correct) and Evidence Ratio (ER, evidence in favor of the effect observed against alternative hypotheses). When ER was weak (below 10, that is, ten times as much evidence for the effect as for alternative hypotheses), we also calculated the ER in favor of the null hypothesis. Note that given the standardization of the outcome variables, the effect size is equivalent to Hedges' g, that is, is expressed in units of standard deviations. To assess the effects of using previous literature informed and skeptical priors, we report the same model estimates for both models. Further, we adopted a Leave-One-Out model comparison framework estimating the model's out-of-sample performance, in other words, estimating the ability of the model to generalize to new data (. We then calculated their relative stacking weight based on Leave-One-Out Information Criteria, assessing the probability of each model to be better than the others (Yao et al., 2018). This procedure informs us as to whether adding information from previous findings in our statistical models enabled us to create more robust models, with greater chances of having replicable findings in new studies.

**Analysis on the association between clinical symptoms and coherence measures**

For ease of comparison with the previous literature reporting Pearson correlation coefficients, we scaled the coherence measures separately for each language, that is, we first subtracted the minimum values and divided the result by range, i.e., the difference between the maximum and minimum values, to bring all values into the zero to one range. We chose not to standardize the clinical features as z-scores, as that would assume a linear relation between coherence measures and clinical features. Clinical features are more adequately modeled as ordinal variables, where a monotonic relation is assumed but non-linear forms are possible. In other words, while we expect that if there is a change in coherence measures when moving from a SANS score of 0 to a score of 1, we should see a change in the same direction when moving from 1 to 2, but the size of the change might be different. Note that in case of linear changes, the current model gives comparable estimates to more traditional models (Bürkner & Charpentier, 2020).

This analysis was performed on the group of patients with schizophrenia only. Indeed, even if clinical ratings were available for many of the HCs, the SANS-SAPS/PANSS scales are designed specifically for assessing symptomatology in individuals with neuropsychiatric disorders and not for capturing individual variability in healthy individuals. As a result, healthy controls present a median score of 0 for the different ratings and minimal variability in scores, which potentially introduces a bias into the model.

Skeptical priors were specified as normal distribution centered at zero (no association between acoustic features and clinical ratings), and a standard deviation of .3. The expected estimates were thus predominantly included within -0.9 and 0.9, but could still be swayed by the data. Intercepts were specified as normal distribution centered at 0.5 and a standard deviation of 0.3. Individual variability was modeled using a positive half-normal prior centered at 0, with a standard deviation of 0.1 from the estimate for the specific group and language of the individual, thus regularizing the inference.

Note that model quality checks (done before actually assessing the results, see Model fitting section and see Figure **SM5_*E*, SM5_*F*, SM5_*G*, SM5_*H***), in particular prior posterior comparisons, indicated that informed priors were too confident in their expectations and strongly limited learning from data. We therefore increased uncertainty by multiplying the standard deviation by 3, upon which the model was better able to also learn from the data, the variance in the posteriors being much lower than in the priors[28]. We then compared results across the informed and skeptical models following the same procedure described in the previous paragraphs.

**Figure SM5_E**. Plot of the prior predictive checks (light blue) and observed data (bold blue) for the relationship between symptoms (SANS Global score) and coherence measures (Coherence 5). Note that the three language groups (DK, GE, CH) are given the same prior expectations for all parameters, thus implicitly assuming no difference between them (so that any evidence for a difference that we might find would come exclusively from the data and not from the priors).

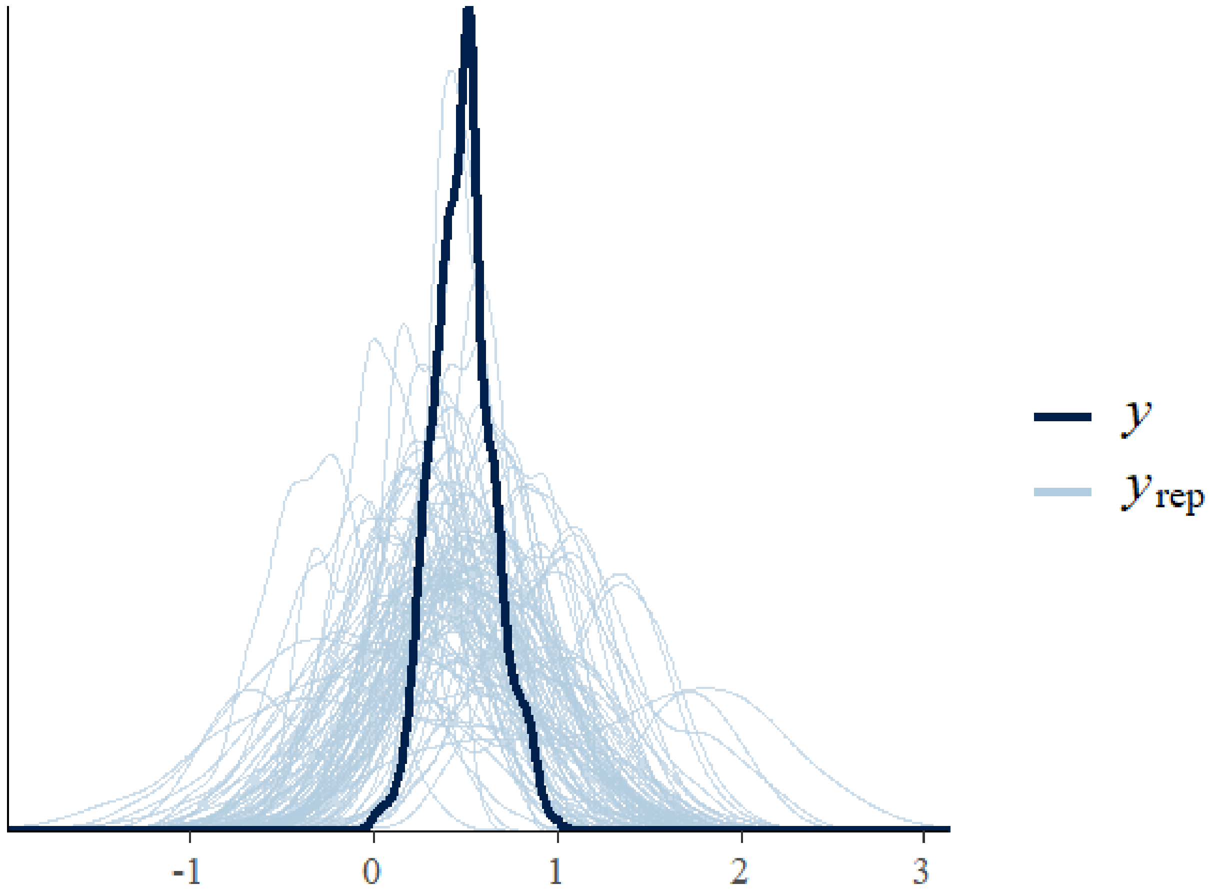

**FIGURE SM5_F.** Plot of the prior-posterior predictive check. Prior (light blue) and posterior (dark blue) distributions *for the intercept* in the model *for the Danish language* for the relationship between symptoms (SANS Global score) and coherence measures (Coherence 5).

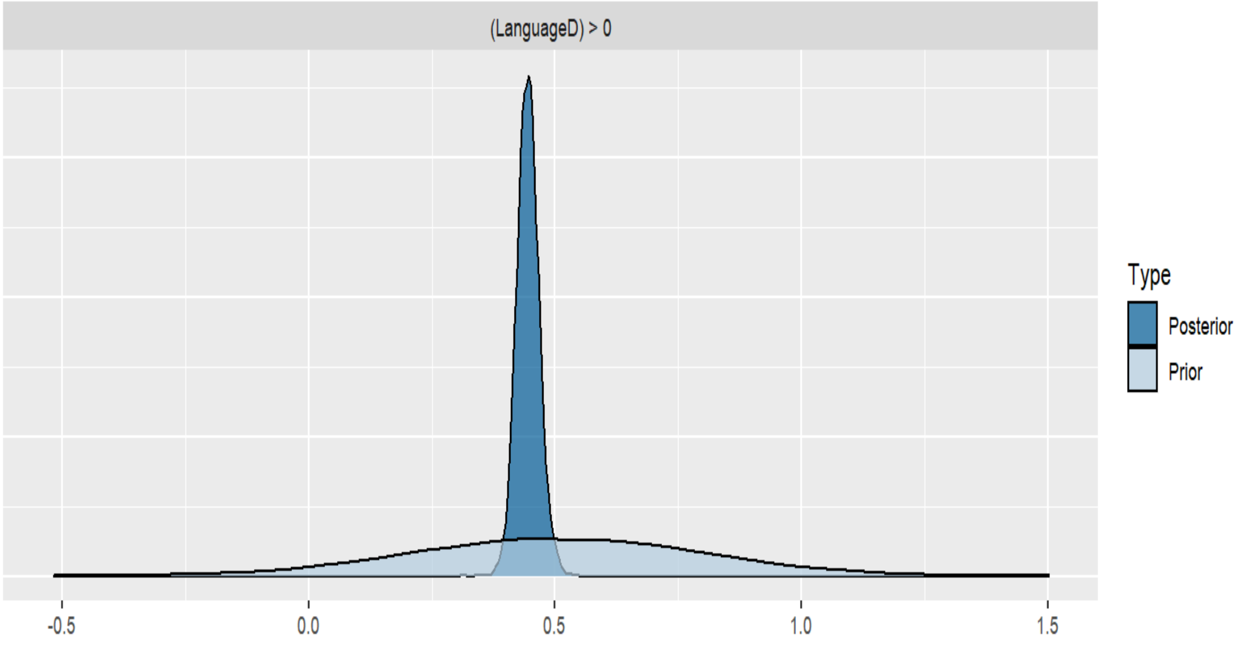


**FIGURE SM5_G.** Plot of the prior-*posterior predictive check.* Prior (light blue) and posterior (dark blue) distributions for the effect of diagnosis (SCZ-HC) for the Danish language for the relationship between symptoms (SANS Global score) and coherence measures (Coherence 5).

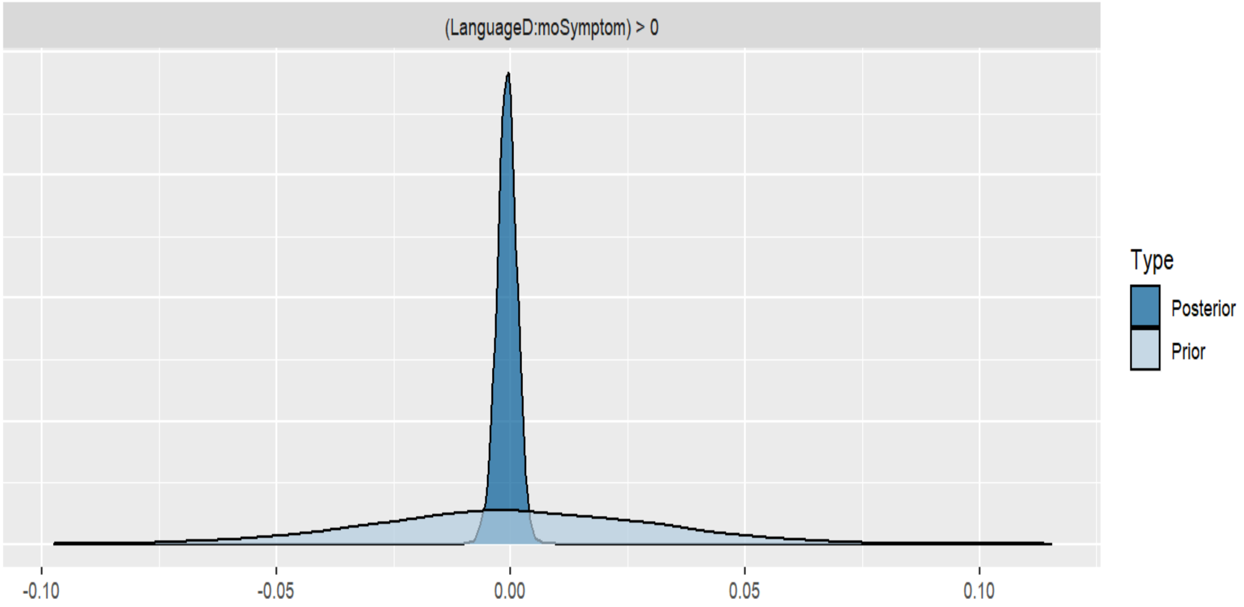

**Figure SM5_H.** Plot of the posterior predictive check. We overlay the distribution of the actual data for the relationship between symptoms (SANS Global score) and coherence measures (Coherence 5) with the distribution of 100 simulations from the fitted model (predictions, each simulation a pale blue line). This allows us to identify potential biases in the model.

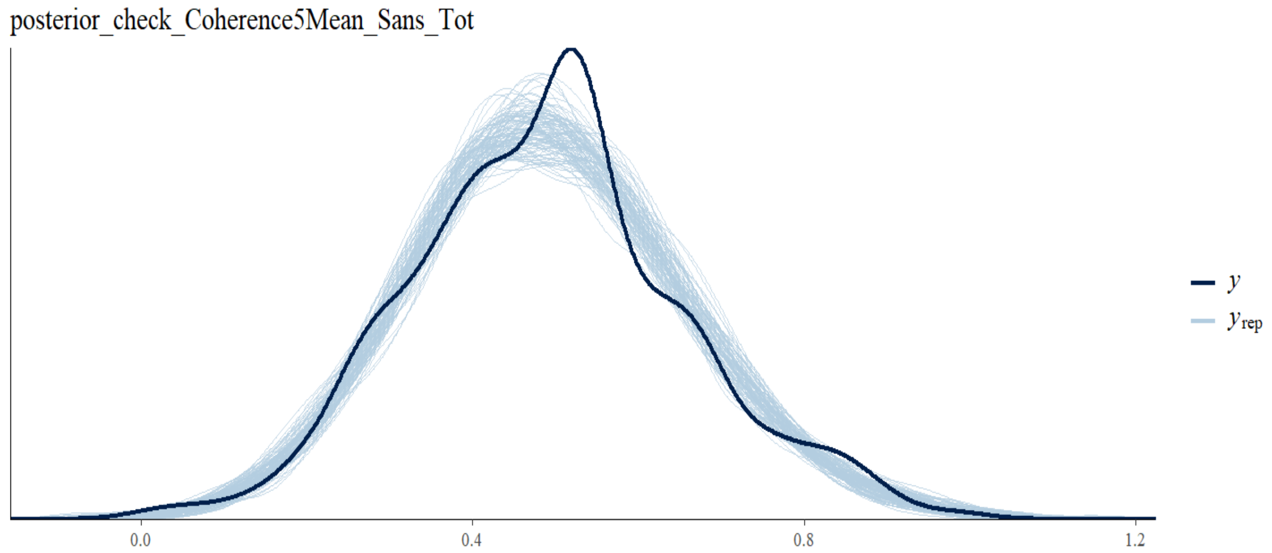


### Results for IQR

The detailed results are reported in **Table SM5_A**. None of the coherence measures robustly generalized across the different languages. Instead, in line with the results for median (see **Results** section and **Table 3** in the main manuscript), we found several specific language effects: in the Chinese corpus we found lower coherence variability in Similarity mean, Coherence 5-10 and Coherence-k measures; in the Danish corpus, we found a mixed pattern with higher coherence variability in Coherence-10 measure, and lower variability in Coherence-k measures, as well as in the German corpus we found lower Coherence 5-10 and Coherence-k variability, and higher First and Second order Coherence variability. Globally, we found important differences within (i.e., between the diverse coherence measures) and between languages.

**Table SM5_A.** Estimated standardized mean difference (HC - patients with schizophrenia) for the fourteen coherence measures, as estimated separately by informed and the skeptical models

| Coherence measure | Group (HC – SCZ) |
|---|---|
| **Similarity Mean** | -0.83 (-1.11 -0.54) |
| Skeptical DK | -0.06 (-0.16 0.05) ER = 4.88 |
| Skeptical GE | -0.04 (-0.22 0.13) ER = 2 ER01 = 4.69 |
| Skeptical CH | **-0.39 (-0.57 -0.21) ER = 1999** |
| **Coherence 5** | |

| | |
|---|---|
| Skeptical DK | 0 (-0.1 0.11) ER = 1.12 ER01 = 7.83 |
| Skeptical GE | **-0.19 (-0.35 -0.02) ER = 28.33** |
| Skeptical CH | **-0.37 (-0.53 -0.2) ER = 3332.33** |
| **Coherence 10** | |
| Skeptical DK | **0.15 (0.06 0.24) ER = 255.41** |
| Skeptical GE | **-0.29 (-0.45 -0.13) ER = 284.71** |
| Skeptical CH | **-0.3 (-0.47 -0.13) ER = 453.55** |
| **Coherence K2** | |
| Skeptical DK | **-0.11 (-0.21 0) ER = 18.57** |
| Skeptical GE | -0.09 (-0.28 0.1) ER = 3.76 |
| Skeptical CH | **-0.22 (-0.38 -0.06) ER = 77.74** |
| **Coherence K3** | |
| Skeptical DK | **-0.1 (-0.2 0) ER = 17.76** |
| Skeptical GE | -0.1 (-0.28 0.08) ER = 4.52 |
| Skeptical CH | **-0.25 (-0.42 -0.08) ER = 143.93** |
| **Coherence K4** | |
| Skeptical DK | -0.04 (-0.12 0.05) ER = 2.99 ER01 = 7.49 |
| Skeptical GE | -0.03 (-0.18 0.12) ER = 1.69 ER01 = 5.38 |
| Skeptical CH | **-0.19 (-0.34 -0.03) ER = 46.62** |
| **Coherence K5** | |
| Skeptical DK | -0.01 (-0.11 0.08) ER = 1.47 ER01 = 8.52 |
| Skeptical GE | -0.11 (-0.3 0.07) ER = 5.28 |
| Skeptical CH | -0.06 (-0.19 0.07) ER = 3.77 |
| **Coherence K6** | |
| Skeptical DK | -0.03 (-0.12 0.05) ER = 2.87 ER01 = 8.03 |
| Skeptical GE | -0.1 (-0.25 0.05) ER = 6.17 |
| Skeptical CH | **-0.12 (-0.25 0.01) ER = 13.04** |
| **Coherence K7** | |
| Skeptical DK | -0.05 (-0.14 0.05) ER = 3.73 |
| Skeptical GE | 0.03 (-0.13 0.2) ER = 1.76 ER01 = 4.97 |
| Skeptical CH | **-0.14 (-0.29 0.01) ER = 16.83** |
| **Coherence K8** | |
| Skeptical DK | -0.02 (-0.11 0.08) ER = 1.5 ER01 = 8.75 |
| Skeptical GE | **-0.23 (-0.38 -0.08) ER = 155.25** |
| Skeptical CH | -0.05 (-0.19 0.08) ER = 2.75 ER01 = 5.29 |
| **Coherence K9** | |
| Skeptical DK | -0.03 (-0.12 0.06) ER = 2.51 ER01 = 7.8 |
| Skeptical GE | **-0.14 (-0.29 0.02) ER = 12.48** |
| Skeptical CH | **-0.28 (-0.43 -0.14) ER = 1110.11** |
| **Coherence K10** | |
| Skeptical DK | -0.05 (-0.14 0.04) ER = 4.32 |
| Skeptical GE | -0.11 (-0.3 0.08) ER = 5.21 |
| Skeptical CH | **-0.29 (-0.43 -0.14) ER = 1110.11** |
| **Coherence first-order** | |
| Skeptical DK | 0.04 (-0.09 0.18) ER = 2.35 ER01 = 5.31 |
| Skeptical GE | **0.21 (0 0.43) ER = 18.46** |
| Skeptical CH | 0.01 (-0.15 0.17) ER = 1.13 ER01 = 5.12 |
| **Coherence second-order** | |
| Skeptical DK | 0.08 (-0.08 0.25) ER = 3.84 |

| | |
|---|---|
| Skeptical GE | **0.26 (-0.02 0.54) ER = 15.21** |
| Skeptical CH | -0.07 (-0.26 0.11) ER = 2.9 ER01 = 3.55 |

### Association between coherence measures and symptoms (PANSS clinical ratings)

**Table SM5_B.** Estimated standardized relation between coherence measures and clinical features (PANSS). ER indicates the evidence ratio for the difference, ER01 the evidence ratio for the null effect.

| Rating scales | PANSS Total | PANSS Negative | PANSS Positive | PANSS Disorganization | PANSS Unusual Thought Content |
|---|---|---|---|---|---|
| **Mean Similarity** | | | | | |
| Skeptical CH | -0.04 (-0.28 0.19) ER = 1.63 ER01 = 4.35 | 0.07 (-0.05 0.21) ER = 4.54 | 0 (-0.15 0.16) ER = 1.08 ER01 = 6.67 | **0.11 (0.02 0.2) ER = 31.97** | -0.06 (-0.18 0.03) ER = 6.33 |
| Skeptical GE | 0.04 (-0.13 0.22) ER = 1.86 ER01 = 5.04 | -0.06 (-0.2 0.07) ER = 3.49 | 0.14 (-0.04 0.36) ER = 8.27 | 0.08 (-0.05 0.3) ER = 5.38 | 0.07 (-0.04 0.21) ER = 5.58 |
| **Coherence 5** | | | | | |
| Skeptical CH | -0.09 (-0.46 0.28) ER = 1.92 ER01 = 2.42 | 0 (-0.22 0.23) ER = 0.98 ER01 = 4.31 | 0.03 (-0.23 0.28) ER = 1.36 ER01 = 3.92 | 0.12 (-0.05 0.29) ER = 7.26 | -0.01 (-0.18 0.17) ER = 1.19 ER01 = 6.44 |
| Skeptical GE | -0.03 (-0.2 0.13) ER = 1.67 ER01 = 5.86 | -0.08 (-0.21 0.03) ER = 7.37 | 0.06 (-0.12 0.24) ER = 2.41 ER01 = 5.5 | 0.05 (-0.09 0.27) ER = 2.58 ER01 = 8.37 | 0.05 (-0.06 0.18) ER = 3.36 |
| **Coherence 10** | | | | | |
| Skeptical CH | -0.14 (-0.56 0.28) ER = 2.3 ER01 = 2.1 | -0.03 (-0.31 0.23) ER = 1.42 ER01 = 3.79 | 0 (-0.32 0.3) ER = 1.01 ER01 = 3.35 | 0.11 (-0.1 0.32) ER = 4.24 | -0.03 (-0.24 0.18) ER = 1.41 ER01 = 5.05 |
| Skeptical GE | -0.01 (-0.21 0.18) ER = 1.17 ER01 = 5.27 | -0.07 (-0.22 0.08) ER = 3.39 | 0.1 (-0.1 0.33) ER = 4.28 | 0.07 (-0.09 0.32) ER = 3.26 | 0.04 (-0.11 0.18) ER = 2.12 ER01 = 6.45 |
| **Coherence k5** | | | | | |
| Skeptical CH | -0.03 (-0.2 0.14) ER = 1.62 ER01 = 5.79 | -0.01 (-0.11 0.08) ER = 1.39 ER01 = 10.12 | -0.01 (-0.12 0.11) ER = 1.12 ER01 = 9.33 | 0.02 (-0.05 0.09) ER = 2.47 ER01 = 12.18 | 0 (-0.08 0.07) ER = 1.1 ER01 = 14.02 |
| Skeptical GE | 0.05 (-0.09 0.2) ER = 2.79 ER01 = 6.2 | -0.01 (-0.12 0.11) ER = 1.17 ER01 = 9.24 | 0.09 (-0.06 0.26) ER = 5.09 | 0.1 (-0.03 0.38) ER = 6.7 | 0.04 (-0.06 0.15) ER = 2.65 ER01 = 9.44 |
| **Coherence k6** | | | | | |
| Skeptical CH | -0.08 (-0.31 0.15) ER = 2.53 ER01 = 4.01 | -0.06 (-0.18 0.07) ER = 3.35 | -0.04 (-0.2 0.11) ER = 2.1 ER01 = 6.54 | 0.03 (-0.08 0.12) ER = 2.2 ER01 = 8.54 | 0 (-0.11 0.1) ER = 0.92 ER01 = 11.04 |
| Skeptical GE | 0 (-0.14 0.14) ER = 1 ER01 = 7.51 | -0.03 (-0.14 0.07) ER = 2.46 ER01 = 8.51 | 0.11 (-0.03 0.27) ER = 9.26 | 0.02 (-0.11 0.19) ER = 1.66 ER01 = 11.25 | 0.01 (-0.09 0.11) ER = 1.31 ER01 = 11.21 |
| **Coherence k7** | | | | | |
| Skeptical CH | **-0.18 (-0.36 -0.02) ER = 33.48** | -0.06 (-0.16 0.02) ER = 7.13 | -0.07 (-0.19 0.04) ER = 5.94 | 0 (-0.08 0.07) ER = 1.06 ER01 = 14.02 | -0.03 (-0.12 0.05) ER = 2.53 ER01 = 12.35 |
| Skeptical GE | 0.1 (-0.02 0.23) ER = 10.74 | 0.05 (-0.05 0.16) ER = 4.06 | 0.09 (-0.04 0.23) ER = 6.77 | 0.1 (-0.01 0.35) ER = 10.32 | 0.06 (-0.03 0.17) ER = 6.19 |

| | | | | | |
|---|---|---|---|---|---|
| **Coherence k8** | | | | | |
| Skeptical CH | **-0.14 (-0.3 0.02) ER = 13.29** | -0.01 (-0.1 0.08) ER = 1.34 ER01 = 11.22 | **-0.09 (-0.2 0.01) ER = 11.27** | 0.01 (-0.06 0.08) ER = 1.41 ER01 = 14.75 | **-0.05 (-0.14 0.01) ER = 10.12** |
| Skeptical GE | 0.02 (-0.08 0.12) ER = 1.73 ER01 = 9.26 | 0.03 (-0.05 0.12) ER = 3.06 | 0.01 (-0.1 0.13) ER = 1.34 ER01 = 9.11 | 0.04 (-0.05 0.23) ER = 2.84 ER01 = 12.73 | -0.02 (-0.1 0.06) ER = 1.56 ER01 = 14.25 |
| **First-order coherence** | | | | | |
| Skeptical CH | -0.01 (-0.3 0.28) ER = 1.14 ER01 = 3.51 | 0.09 (-0.08 0.26) ER = 4.06 | -0.09 (-0.29 0.1) ER = 3.79 | -0.08 (-0.2 0.05) ER = 6.04 | -0.01 (-0.14 0.13) ER = 1.33 ER01 = 7.66 |
| Skeptical GE | -0.13 (-0.49 0.26) ER = 2.46 ER01 = 2.24 | -0.14 (-0.42 0.17) ER = 3.82 | 0.02 (-0.37 0.42) ER = 1.13 ER01 = 2.59 | -0.11 (-0.48 0.18) ER = 3.05 | -0.07 (-0.35 0.22) ER = 2.26 ER01 = 3.23 |
| **Second-order coherence** | | | | | |
| Skeptical CH | 0.06 (-0.28 0.4) ER = 1.62 ER01 = 2.83 | 0.09 (-0.12 0.29) ER = 3.27 | 0.05 (-0.19 0.29) ER = 1.65 ER01 = 4.25 | 0.06 (-0.1 0.22) ER = 3.19 | 0.01 (-0.17 0.18) ER = 1.09 ER01 = 6.36 |
| Skeptical GE | -0.25 (-0.61 0.1) ER = 7.23 | -0.06 (-0.33 0.21) ER = 1.81 ER01 = 3.57 | -0.14 (-0.48 0.18) ER = 3.44 | -0.19 (-0.56 0.07) ER = 7.31 | -0.16 (-0.41 0.08) ER = 6.94 |
| **Total Words** | | | | | |
| Skeptical CH | **0.2 (0.02 0.38) ER = 25.79** | **0.1 (0 0.2) ER = 18.74** | 0.08 (-0.04 0.21) ER = 6.8 | 0 (-0.08 0.09) ER = 1 ER01 = 11.47 | 0.03 (-0.05 0.11) ER = 2.7 ER01 = 11.19 |
| Skeptical GE | -0.13 (-0.36 0.11) ER = 4.51 | **-0.16 (-0.35 0.02) ER = 14.19** | 0.11 (-0.15 0.38) ER = 3.32 | 0 (-0.24 0.23) ER = 1.02 ER01 = 6.7 | 0.02 (-0.15 0.21) ER = 1.36 ER01 = 6.32 |

### SM6 – Robustness analysis

In the analyses reported in the main manuscript, fillers such as 'uhm' and 'ehm' were removed before deriving the various coherence measures. As suggested by Iter et al., 2018 (see also Elvevåg et al., 2007), coherence measures can be biased by: 1) verbal fillers, (2) longer sentences and (3) repetitions. The authors indeed indicated how the class of algorithms which use cosine similarity to model semantic similarity and concept overlap may incorrectly attribute higher coherence to longer sentences and greater use of verbal fillers and repetitions. In this robustness analysis, we replicated the analysis described in the main manuscript by: 1) including verbal fillers 2) including verbal fillers and punctuations; 3) we also explicitly modeled the relationship between transcript length (total number of words) and the various coherence measures. The aim of this procedure was to assess whether our analysis is robust to different preprocessing methods and the relative impact of each of these components (i.e., fillers, punctuation and transcript length) on the results. The results are reported in **Table SM6** and qualitative comparisons with the results described in the main manuscript (see section Results) are reported below.

#### Differences between the different cleaning options

We found no differences in the Danish corpus: results were robust to changes in the cleaning options. Instead, we found differences in the German corpus once we changed the cleaning options. In particular, including punctuations and fillers in the analysis, we found that the differences between patients with schizophrenia and controls in some coherence measures (Coherence 10, Coherence-k, Second-order coherence) varied. For the Chinese corpus, we found some variation in the Coherence-k and Second-order coherence when including punctuations and fillers in the analysis, but results were generally robust to changes in the cleaning options. Overall, these results show how NLP measures of coherence, and in particular cosine similarity derived measures, are sensitive to fillers and punctuations, and how these components can introduce bias in the analysis. As for transcript length, in line with previous literature (Elvevag et al., 2007; Iter et al., 2018; Corcoran et al., 2018; Hitczenko et al., 2022) we found that transcript length is positively associated with the various coherence measures, that is longer transcripts have generally higher coherence values. This confirms that the impact of transcript length should be controlled in the analysis in order to avoid possible bias in the results.

**Table SM6**. Comparison of the estimated standardized mean difference (HC - patients with schizophrenia) for the 14 coherence measures, as estimated separately by informed and the skeptical models, between the different cleaning options and relationship between coherence measures and transcript length (total number of words).

| **Coherence measure** | **No cleaning** | **No fillers** | **No fillers and no punctuation** | **Length** |
|---|---|---|---|---|
| | **Group (CT – SCZ)** | **Group (CT – SCZ)** | **Group (CT – SCZ)** | |
| **Similarity Mean** | -0.83 (-1.11 -0.54) | -0.83 (-1.11 -0.54) | -0.83 (-1.11 -0.54) | |
| Skeptical DK | -0.01 (-0.12 0.1) ER = 1.32 ER01 = 7.37 | 0.01 (-0.1 0.12) ER = 1.27 ER01 = 6.79 | 0.04 (-0.06 0.15) ER = 2.9 ER01 = 6.44 | **0.15 (0.09 0.21) ER = Inf** |
| Skeptical GE | -0.07 (-0.23 0.09) ER = 3.25 | 0.04 (-0.12 0.19) ER = 1.91 ER01 = 4.78 | -0.03 (-0.19 0.12) ER = 1.76 ER01 = 5.3 | 0.03 (-0.06 0.13) ER = 2.41 ER01 = 7.79 |
| Skeptical CH | **-0.38 (-0.58 -0.17) ER = 665.67** | **-0.42 (-0.62 -0.21) ER = 908.09** | **-0.27 (-0.47 -0.07) ER = 77.12** | 0.03 (-0.07 0.14) ER = 2.3 ER01 = 6.95 |
| Informed DK | **-0.15 (-0.26 -0.05) ER = 113.94** | **-0.14 (-0.24 -0.03) ER = 73.07** | **-0.1 (-0.2 0) ER = 16.7** | NA |
| Informed GE | **-0.32 (-0.48 -0.17) ER = Inf** | **-0.25 (-0.41 -0.09) ER = 276.78** | **-0.29 (-0.45 -0.14) ER = 2499** | NA |
| Informed CH | **-0.59 (-0.75 -0.43) ER = Inf** | **-0.61 (-0.76 -0.45) ER = Inf** | **-0.52 (-0.68 -0.36) ER = Inf** | NA |
| *Stacking weight* | **Skeptic Model 0.820** | **Skeptic Model 0.82** | **Skeptic Model 0.93** | |
| **Coherence 5** | -0.26 (-0.58 0.06) | -0.26 (-0.58 0.06) | -0.26 (-0.58 0.06) | |
| Skeptical DK | 0.06 (-0.06 0.18) ER = 3.94 | 0.07 (-0.07 0.2) ER = 3.94 | 0.09 (-0.03 0.21) ER = 9.45 | **0.24 (0.19 0.31) ER = Inf** |
| Skeptical GE | -0.06 (-0.23 0.11) ER = 2.48 ER01 = 4.24 | 0.05 (-0.12 0.22) ER = 2.17 ER01 = 4.51 | -0.13 (-0.3 0.05) ER = 7.84 | 0.03 (-0.06 0.12) ER = 2.39 ER01 = 7.9 |
| Skeptical CH | **-0.33 (-0.55 -0.12) ER = 157.73** | **-0.35 (-0.57 -0.13) ER = 249** | **-0.41 (-0.62 -0.19) ER = 665.67** | **0.13 (0.03 0.23) ER = 49** |
| Informed DK | 0.01 (-0.11 0.12) ER = 1.21 ER01 = 7.9 | 0 (-0.11 0.12) ER = 1.07 ER01 = 7.91 | 0.03 (-0.08 0.14) ER = 2.2 ER01 = 7.44 | NA |
| Informed GE | -0.12 (-0.27 0.03) ER = 9.62 | -0.05 (-0.19 0.1) ER = 2.26 ER01 = 6.41 | **-0.17 (-0.32 -0.03) ER = 41.19** | NA |
| Informed CH | **-0.32 (-0.49 -0.14) ER = 624** | **-0.33 (-0.5 -0.16) ER = 999** | **-0.36 (-0.53 -0.2) ER = 3332.33** | NA |
| *Stacking weight* | **Informed Model 1.0** | **Skeptic Model 0.99** | **Skeptic Model 0.77** | |

| | | | | |
|---|---|---|---|---|
| **Coherence 10** | -0.09 (-0.41 0.23) | -0.09 (-0.41 0.23) | -0.09 (-0.41 0.23) | |
| Skeptical DK | **0.11 (-0.02 0.23) ER = 11.48** | **0.15 (0.02 0.28) ER = 32.67** | **0.15 (0.03 0.28) ER = 41.02** | **0.23 (0.17 0.29) ER = Inf** |
| Skeptical GE | -0.06 (-0.24 0.14) ER = 2.17 ER01 = 3.93 | 0.05 (-0.14 0.23) ER = 1.89 ER01 = 4.16 | **-0.2 (-0.39 -0.01) ER = 21.57** | 0.03 (-0.06 0.12) ER = 2.23 ER01 = 8.2 |
| Skeptical CH | **-0.37 (-0.58 -0.15) ER = 249** | **-0.38 (-0.59 -0.17) ER = 713.29** | **-0.42 (-0.65 -0.2) ER = 1665.67** | **0.11 (0.01 0.21) ER = 29.3** |
| Informed DK | 0.07 (-0.04 0.19) ER = 5.74 | **0.1 (-0.02 0.22) ER = 10.31** | **0.11 (0 0.22) ER = 17.02** | NA |
| Informed GE | -0.07 (-0.22 0.09) ER = 3.18 | 0 (-0.15 0.15) ER = 1.02 ER01 = 2.14 | **-0.17 (-0.33 -0.01) ER = 23.45** | NA |
| Informed CH | **-0.27 (-0.44 -0.09) ER = 155.25** | **-0.28 (-0.45 -0.11) ER = 242.9** | **-0.3 (-0.47 -0.13) ER = 453.55** | NA |
| *Stacking weight* | **Informed Model 0.99** | **Skeptic Model 1.0** | **Skeptic Model 0.5** | |
| **Coherence K2** | -0.33 (-0.88 0.23) | -0.33 (-0.88 0.23) | -0.33 (-0.88 0.23) | |
| Skeptical DK | 0.08 (-0.04 0.19) ER = 6.76 | **0.13 (0.01 0.25) ER = 24.91** | **0.16 (0.04 0.27) ER = 87.5** | **0.11 (0.05 0.17) ER = 1110.11** |
| Skeptical GE | -0.11 (-0.29 0.07) ER = 5.25 | -0.04 (-0.21 0.13) ER = 1.83 ER01 = 4.66 | -0.12 (-0.27 0.04) ER = 8.09 | **-0.13 (-0.22 -0.04) ER = 134.14** |
| Skeptical CH | **-0.21 (-0.42 -0.01) ER = 22.53** | **-0.23 (-0.44 -0.03) ER = 29.96** | -0.16 (-0.4 0.09) ER = 5.61 | 0.04 (-0.06 0.14) ER = 2.77 ER01 = 6.36 |
| Informed DK | 0.06 (-0.05 0.16) ER = 3.81 | **0.1 (-0.02 0.21) ER = 11.38** | **0.13 (0.02 0.24) ER = 41.74** | NA |
| Informed GE | **-0.14 (-0.31 0.02) ER = 11.72** | -0.08 (-0.24 0.08) ER = 3.68 | **-0.15 (-0.3 0) ER = 17.45** | NA |
| Informed CH | **-0.24 (-0.44 -0.05) ER = 52.76** | **-0.26 (-0.45 -0.07) ER = 75.34** | **-0.2 (-0.42 0.02) ER = 14.58** | NA |
| | **Skeptic Model 1.0** | **Skeptic Model 1.0** | **Skeptic Model 1.0** | |
| **Coherence K3** | -0.19 (-0.74 0.36) | -0.19 (-0.74 0.36) | -0.19 (-0.74 0.36) | |
| Skeptical DK | **0.15 (0.04 0.26) ER = 69.92** | **0.17 (0.05 0.28) ER = 124** | **0.11 (-0.01 0.22) ER = 14.77** | **0.18 (0.12 0.24) ER = Inf** |
| Skeptical GE | 0 (-0.17 0.16) ER = 1.02 ER01 = 4.88 | 0.09 (-0.08 0.25) ER = 4.31 | **-0.2 (-0.36 -0.03) ER = 37.91** | 0.03 (-0.06 0.12) ER = 2.18 ER01 = 7.87 |
| Skeptical CH | -0.13 (-0.36 0.1) ER = 4.73 | -0.14 (-0.37 0.09) ER = 4.95 | **-0.31 (-0.51 -0.11) ER = 195.08** | 0.07 (-0.03 0.18) ER = 7.05 |
| Informed DK | **0.13 (0.02 0.24) ER = 43.64** | **0.15 (0.04 0.26) ER = 82.33** | 0.09 (-0.02 0.2) ER = 9.4 | NA |
| Informed GE | -0.02 (-0.18 0.13) ER = 1.51 ER01 = 3.73 | 0.06 (-0.11 0.22) ER = 2.59 ER01 = 3.03 | **-0.2 (-0.36 -0.04) ER = 58.88** | NA |
| Informed CH | -0.15 (-0.35 0.06) ER = 7.54 | -0.15 (-0.36 0.05) ER = 8.02 | **-0.31 (-0.5 -0.12) ER = 242.9** | NA |
| | **Informed Model 1.0** | **Skeptic Model 1.000** | **Informed Model 1.0** | |
| **Coherence K4** | -0.18 (-0.73 0.37) | -0.18 (-0.73 0.37) | -0.18 (-0.73 0.37) | |
| Skeptical DK | **0.08 (-0.01 0.18) ER = 15.53** | **0.14 (0.04 0.24) ER = 98.01** | **0.14 (0.04 0.24) ER = 112.64** | **0.12 (0.07 0.18) ER = Inf** |
| Skeptical GE | 0.12 (-0.04 0.28) ER = 7.63 | **0.19 (0.03 0.34) ER = 38.06** | -0.05 (-0.21 0.11) ER = 2.35 ER01 = 4.52 | -0.05 (-0.15 0.04) ER = 4.73 |
| Skeptical CH | **-0.27 (-0.47 -0.07) ER = 67.97** | **-0.28 (-0.48 -0.08) ER = 100.01** | **-0.4 (-0.6 -0.21) ER = 1249** | **0.09 (-0.01 0.2) ER = 12.09** |
| Informed DK | 0.08 (-0.01 0.17) ER = 12.07 | **0.13 (0.03 0.23) ER = 65.23** | **0.13 (0.04 0.22) ER = 91.59** | NA |
| Informed GE | 0.09 (-0.06 0.24) ER = 5.19 | **0.15 (0.01 0.3) ER = 22.31** | -0.07 (-0.22 0.09) ER = 3.11 | NA |
| Informed CH | -0.27 (-0.46 -0.08) ER = 101.04 | **-0.28 (-0.46 -0.09) ER = 132.33** | **-0.39 (-0.57 -0.2) ER = 1249** | NA |
| | **Skeptic Model 1.0** | **Skeptic Model 1.000** | **Informed Model 1.0** | |

| | | | | |
|---|---|---|---|---|
| **Coherence K5** | -0.18 (-0.73 0.37) | -0.18 (-0.73 0.37) | -0.18 (-0.73 0.37) | |
| Skeptical DK | **0.15 (0.06 0.23) ER = 195.08** | **0.2 (0.1 0.3) ER = 2499** | **0.17 (0.08 0.27) ER = 356.14** | **0.09 (0.03 0.15) ER = 174.44** |
| Skeptical GE | 0.05 (-0.12 0.21) ER = 2.29 ER01 = 4.36 | **0.13 (-0.03 0.29) ER = 9.95** | -0.1 (-0.27 0.07) ER = 4.98 | 0.03 (-0.07 0.13) ER = 2.26 ER01 = 7.83 |
| Skeptical CH | **-0.15 (-0.33 0.02) ER = 12.62** | **-0.15 (-0.32 0.01) ER = 14.48** | **-0.25 (-0.4 -0.1) ER = 262.16** | 0.07 (-0.03 0.17) ER = 7.49 |
| Informed DK | **0.13 (0.04 0.22) ER = 162.93** | **0.18 (0.09 0.28) ER = 499** | **0.16 (0.06 0.26) ER = 262.16** | NA |
| Informed GE | 0.03 (-0.13 0.18) ER = 1.59 ER01 = 3.56 | 0.1 (-0.05 0.25) ER = 6.05 | -0.12 (-0.28 0.05) ER = 7.5 | NA |
| Informed CH | **-0.16 (-0.33 0) ER = 17.69** | **-0.16 (-0.32 0) ER = 18.34** | **-0.25 (-0.39 -0.1) ER = 453.55** | NA |
| | **Skeptic Model 1.000** | **Informed Model 0.65** | **Informed Model 0.88** | |
| **Coherence K6** | | | | |
| Skeptical DK | **0.14 (0.05 0.24) ER = 152.85** | **0.16 (0.05 0.26) ER = 130.58** | 0.07 (-0.03 0.16) ER = 7.16 | **0.11 (0.05 0.17) ER = 999** |
| Skeptical GE | 0.05 (-0.13 0.23) ER = 2.26 ER01 = 4.14 | 0.09 (-0.1 0.28) ER = 3.91 | **-0.18 (-0.35 0) ER = 18.08** | 0 (-0.1 0.09) ER = 1.07 ER01 = 8.93 |
| Skeptical CH | **-0.24 (-0.41 -0.07) ER = 82.33** | **-0.28 (-0.45 -0.11) ER = 269.27** | **-0.27 (-0.45 -0.09) ER = 165.67** | 0.05 (-0.06 0.15) ER = 3.57 |
| **Coherence K7** | | | | |
| Skeptical DK | **0.14 (0.04 0.24) ER = 87.5** | **0.13 (0.03 0.23) ER = 57.82** | **0.11 (0.01 0.22) ER = 26.25** | **0.13 (0.07 0.19) ER = 1999** |
| Skeptical GE | 0.04 (-0.12 0.2) ER = 1.93 ER01 = 4.5 | 0.1 (-0.05 0.26) ER = 6.59 | -0.06 (-0.22 0.11) ER = 2.55 ER01 = 4.42 | -0.06 (-0.16 0.03) ER = 6.77 |
| Skeptical CH | **-0.36 (-0.55 -0.17) ER = 832.33** | **-0.34 (-0.52 -0.15) ER = 554.56** | **-0.32 (-0.5 -0.13) ER = 262.16** | **0.1 (0 0.19) ER = 21.17** |
| **Coherence K8** | | | | |
| Skeptical DK | 0.08 (-0.03 0.18) ER = 8.32 | **0.17 (0.06 0.28) ER = 150.52** | **0.13 (0.02 0.23) ER = 46.39** | **0.15 (0.09 0.21) ER = Inf** |
| Skeptical GE | 0.07 (-0.1 0.24) ER = 3.21 | 0.12 (-0.05 0.3) ER = 7.66 | -0.11 (-0.28 0.06) ER = 5.86 | -0.07 (-0.16 0.02) ER = 9.89 |
| Skeptical CH | **-0.28 (-0.49 -0.07) ER = 69.42** | **-0.31 (-0.52 -0.11) ER = 137.89** | **-0.22 (-0.4 -0.03) ER = 35.5** | 0.08 (-0.03 0.18) ER = 8.67 |
| **Coherence K9** | | -0.18 (-0.73 0.37) | | |
| Skeptical DK | **0.08 (-0.01 0.18) ER = 12.48** | 0.09 (-0.03 0.21) ER = 7.72 | **0.12 (-0.02 0.27) ER = 11.92** | **0.13 (0.06 0.2) ER = 1999** |
| Skeptical GE | -0.04 (-0.2 0.13) ER = 1.77 ER01 = 4.72 | **0.16 (-0.02 0.34) ER = 13.75** | -0.09 (-0.27 0.09) ER = 4.12 | 0 (-0.09 0.09) ER = 1.03 ER01 = 8.85 |
| Skeptical CH | **-0.19 (-0.38 0) ER = 20.6** | **-0.3 (-0.48 -0.12) ER = 249** | **-0.36 (-0.56 -0.17) ER = 768.23** | 0.06 (-0.04 0.16) ER = 4.92 |
| **Coherence K10** | | | | |
| Skeptical DK | 0.09 (-0.02 0.21) ER = 9.74 | **0.13 (0.01 0.24) ER = 24.97** | **0.12 (0.02 0.21) ER = 33.6** | **0.13 (0.07 0.2) ER = 3332.33** |
| Skeptical GE | **0.16 (0 0.31) ER = 18.92** | **0.23 (0.07 0.39) ER = 84.47** | -0.08 (-0.25 0.08) ER = 3.87 | 0.01 (-0.08 0.11) ER = 1.37 ER01 = 8.76 |
| Skeptical CH | **-0.3 (-0.48 -0.11) ER = 187.68** | **-0.3 (-0.48 -0.11) ER = 262.16** | **-0.37 (-0.56 -0.17) ER = 3332.33** | 0.06 (-0.04 0.16) ER = 5 |
| **Coherence first-order** | | -0.19 (-0.9 0.51) | -0.19 (-0.9 0.51) | |
| Skeptical DK | **-0.28 (-0.42 -0.14) ER = 2499** | **-0.27 (-0.41 -0.14) ER = 2499** | **-0.33 (-0.47 -0.2) ER = Inf** | **0.5 (0.44 0.57) ER = Inf** |
| Skeptical GE | 0.11 (-0.17 0.38) ER = 2.94 ER01 = 2.43 | 0.05 (-0.22 0.32) ER = 1.58 ER01 = 2.84 | -0.11 (-0.38 0.17) ER = 3.01 | **0.45 (0.34 0.55) ER = Inf** |
| Skeptical CH | **-0.16 (-0.34 0.02) ER = 13.25** | **-0.16 (-0.33 0.02) ER = 13.62** | **-0.26 (-0.42 -0.09) ER = 157.73** | **0.35 (0.27 0.44) ER = Inf** |

| | | | | |
|---|---|---|---|---|
| Informed DK | **-0.29 (-0.42 -0.15) ER = 4999** | **-0.28 (-0.41 -0.14) ER = 4999** | **-0.34 (-0.46 -0.21) ER = Inf** | NA |
| Informed GE | 0.06 (-0.21 0.32) ER = 1.79 ER01 = 2.47 | 0 (-0.27 0.26) ER = 0.98 ER01 = 2.52 | -0.13 (-0.38 0.12) ER = 4.14 | NA |
| Informed CH | **-0.17 (-0.34 0.01) ER = 17.32** | **-0.17 (-0.34 0) ER = 18.84** | **-0.27 (-0.42 -0.11) ER = 311.5** | NA |
| *Stacking weight* | **Skeptic Model 1.0** | **Skeptic Model 0.98** | **Skeptic Model 0.64** | |
| **Coherence second-order** | | | | |
| Skeptical DK | **-0.15 (-0.32 0.01) ER = 14.22** | **-0.15 (-0.32 0.02) ER = 13.51** | **-0.2 (-0.38 -0.03) ER = 30.85** | **0.48 (0.39 0.56) ER = Inf** |
| Skeptical GE | 0.09 (-0.27 0.46) ER = 2.01 ER01 = 2.06 | -0.04 (-0.38 0.29) ER = 1.4 ER01 = 2.39 | **-0.26 (-0.56 0.04) ER = 12.09** | **0.3 (0.18 0.42) ER = Inf** |
| Skeptical CH | -0.12 (-0.32 0.08) ER = 5.35 | -0.12 (-0.31 0.08) ER = 5.03 | **-0.2 (-0.4 0.01) ER = 17.02** | **0.25 (0.16 0.35) ER = Inf** |
| **Total Words** | | | | |
| Skeptical DK | **-0.21 (-0.36 -0.05) ER = 59.61** | **-0.21 (-0.36 -0.05) ER = 52.19** | **-0.21 (-0.37 -0.05) ER = 80.3** | NA |
| Skeptical GE | 0.1 (-0.18 0.38) ER = 2.62 ER01 = 2.54 | 0.06 (-0.21 0.33) ER = 1.72 ER01 = 2.99 | -0.1 (-0.37 0.18) ER = 2.59 ER01 = 2.52 | NA |
| **Skeptical CH** | **-0.24 (-0.48 0) ER = 19.92** | **-0.24 (-0.47 0) ER = 18.34** | **-0.25 (-0.49 -0.01) ER = 21.17** | NA |

### SM7 - Software implementation notes

Bayesian meta-analyses were calculated using the brms (Bürkner et al., 2017) R interface for Stan (Gelman et al., 2015). Figures were produced with ggplot2 from the tidyverse package (Wickham & Wickham, 2017). All analyses were conducted in R using the RStudio IDE (R Core Team, 2020). The source code for the analysis is openly available and can be found at: https://osf.io/8btp6/ .
